# Parental income and psychiatric disorders from age 10 to 40: a genetically informative population study

**DOI:** 10.1101/2024.10.21.24315865

**Authors:** Hans Fredrik Sunde, Espen Moen Eilertsen, Jonas Minet Kinge, Thomas H. Kleppesto, Magnus Nordmo, Avshalom Caspi, Terrie E. Moffitt, Fartein Ask Torvik

**Affiliations:** Centre for Fertility and Health, Norwegian Institute of Public Health, Oslo, Norway; PsychGen Centre for Genetic Epidemiology and Mental Health, Norwegian Institute of Public Health, Oslo, Norway; Department of Psychology, University of Oslo, Oslo, Norway; PROMENTA Research Centre, Department of Psychology, University of Oslo, Oslo, Norway; Department of Health Management and Health Economics, University of Oslo, Oslo, Norway; Department of Psychology, Norwegian University of Science and Technology, Trondheim, Norway; Department of Educational Science, University of South-Eastern Norway, Notodden, Norway; Institute of Psychiatry, Psychology, & Neuroscience, King’s College London, London, UK; Department of Psychology & Neuroscience, Duke University, Durham, NC, USA

**Keywords:** psychiatric disorders, socioeconomic status, social selection, social causation, children of twins, registry data

## Abstract

**Background:** Lower parental income is associated with more psychiatric disorders among offspring, but it is unclear if this association reflects effects of parental income (social causation) or shared risk factors (social selection). Prior research finds contradictory results, which may be due to age differences between the studied offspring.

**Methods:** Here, we studied psychiatric disorders in the entire Norwegian population aged 10 to 40 between 2006 and 2018 (N = 2,468,503). By linking tax registries to administrative health registries, we described prevalence rates by age, sex, and parental income rank. Next, we grouped observations into age groups (adolescence, ages 10-20; early adulthood, 21-30; adulthood, 30-40) and applied kinship-based models with extended families of twins and siblings to decompose the parent-offspring correlation into phenotypic transmission, passive genetic transmission, and passive environmental transmission.

**Results:** We found that lower parental income rank was associated with higher prevalence of nearly all psychiatric disorders, except for eating disorders, for both men and women at all ages from age 10 to 40. Comparing the top with the bottom paternal income quartile, the prevalence ratio of any psychiatric disorder was 0.47 among 10-year-olds and decreased to 0.72 among 40-year-olds. The parent-offspring correlation was −.15 in adolescence, −.10 in early adulthood, and −.06 in adulthood. The kinship-based models indicated that phenotypic transmission could account for 39% of the parent-offspring correlation among adolescents (*p* < .001), but with no significant contribution in early adulthood (*p =* .181) or adulthood (*p* = .737). Passive genetic and environmental transmission contributed to the parent-offspring correlation in all age groups (all *p*’s < .001).

**Conclusion:** Our findings are consistent with a significant role of social causation during adolescence, while social selection could fully explain the parent-offspring correlation in adulthood.

**Key Points:**

- Lower parental income is associated with more psychiatric disorders among offspring.
- There are conflicting results whether this association reflects effects of parental income or shared risk factors.
- Analysing over 2.4 million individuals, our study provides a comprehensive overview of psychiatric disorder prevalence across age, sex, and parental income rank.
- After accounting for shared genetics and environments, we find that parental income rank appears to be associated with psychiatric disorders in adolescence, but not in adulthood.
- The findings support early interventions targeting socioeconomic disparities to reduce adolescent psychiatric disorders.

---

Adults with lower income experience more health problems, including mental health problems, than their more well-off peers (Bor et al., 2017; Fryers et al., 2003; Hu et al., 2016; Kinge et al., 2019; Meng et al., 2020). This health gradient is reproduced across generations: Children from low-income households are at greater risk for poor health outcomes, including mental health problems (Evensen et al., 2021; Kinge et al., 2021; Sariaslan et al., 2021; Torvik et al., 2020). Despite being well documented, it is not understood how the association between parental income and psychiatric disorders in offspring arises.

There are at least two potential causes (Kröger et al., 2015; Ritsher et al., 2001; Thomson et al., 2022). First, growing up in low-income households can directly increase the risk of psychiatric disorders. This is the *social causation* hypothesis. Alternatively, income could be influenced by psychiatric disorders or risk factors related to psychiatric disorders. This is the *social selection* hypothesis. Here, propensity for psychiatric disorders is correlated with propensity for less income. This propensity may be genetically or environmentally inherited by offspring, thus causing an intergenerational association between parental income and offspring psychiatric disorders.

Earlier attempts at distinguishing social causation and social selection have often focused on explaining within-individual associations, where the evidence points to a non-zero role for social causation (Demange et al., 2024; Kröger et al., 2015; Meng et al., 2020; Ritsher et al., 2001; Thomson et al., 2022). However, within-individual and intergenerational associations need not have the same explanation. For intergenerational associations, the evidence is mixed. Some studies support the social selection hypothesis (Halpern-Manners et al., 2016; Sariaslan et al., 2021; Tambs et al., 2012). For example, Sariaslan et al. (2021) found no evidence for social causation when comparing adult Finnish siblings who had experienced different parental income during childhood. This is corroborated by twin studies of psychiatric disorders, which typically find limited evidence for environmental effects shared by siblings (Polderman et al., 2015). Other studies are suggestive of social causation (Akee et al., 2018; Costello et al., 2003; Kinge et al., 2021; Ritsher et al., 2001). Costello et al. (2003) used a natural experiment to conclude that randomly moving Native American families out of poverty reduced symptoms of behavioural disorders in their children, and Kinge et al. (2021) observed a health gradient along parental income and offspring psychiatric disorders among adoptive children in Norway, although it was less steep than among non-adoptive children. Because the intergenerational association is not confounded by shared genetic propensities for these children, these studies suggest that social causation may play a role in reproducing the health gradient across generations.

The mixed results in the literature can have numerous explanations, ranging from sociopolitical differences between the populations under study, the methods employed (Biele et al., 2022; Keyes & Susser, 2022; Rod et al., 2021), or the ages at which the outcomes are observed. For example, whereas *Kinge et al.* and *Costello et al.* investigated psychiatric disorders in children, *Sariaslan et al.* investigated psychiatric disorders in adults. Perhaps parental income affects children’s mental health when the children are residing with their parents, but the effects diminish as children move out into the world on their own. Environmental effects shared by family members are known to diminish with age for some phenotypes, including psychiatric phenotypes (Bergen et al., 2007; Polderman et al., 2015). Alternatively, methodological differences may explain the discrepancy. For example, Biele et al. (2022) also found null results when comparing child siblings. To reconcile these studies, it is necessary to investigate offspring at different ages and to tackle the causation-selection issue with a diverse range of methodologies.

One promising methodology for tackling the causation-selection issue is kinship-based models of extended families, such as the children-of-twins model (D’Onofrio et al., 2003; Mcadams et al., 2018; Sunde et al., 2024). Kinship-based models employ differences in genetic relatedness to differentiate genetic and environmental causes of correlations between family members. Because monozygotic twins are genetically identical, their offspring will be equally related to their parent and their parent’s twin. If the correlation between parents and offspring was entirely due to genetic similarity, you would expect the avuncular correlation (i.e., the correlation between offspring and their uncle or aunt) in families of monozygotic twins to be the same as the parent-offspring correlation. If parents instead had a causal effect on their offspring, the parent-offspring correlation should be greater than the avuncular correlation. In regular families, where offspring are twice as related to their parents as their uncles and aunts, the avuncular correlation can inform on whether shared risk factors are primarily genetic or environmental in origin. In this study, we aim to accurately describe the prevalence rates of psychiatric disorders for the Norwegian population across age and parental income, and to apply such kinship-based models to understand the cause of the intergenerational health gradient.

## Methods

### Sample

Our sample is based on the population register of Norway, which covers all inhabitants of Norway from 1960 to today (N = 8 589 458). Using unique personal identifiers, it is possible to link together tax records, health records, and basic demographic information on the entire population. The demographic information includes data on parentage for most of the population, which allowed us to link parental income data to offspring health data. The available health records span 13 years from 2006 to 2018. We investigated every individual who was aged between 10 and 40 at some point between 2006 and 2018, with available parental income data, and who was alive and living in Norway at the beginning of the relevant years. Persons who temporarily migrated were not included in years they lived abroad. There were 2,468,503 unique eligible individuals resulting in 22,826,805 person-years of data for the descriptive analyses (approx. 736,000 individuals per age, see Supplementary Table S1 for precise numbers for each age).

For the kinship-based models, we grouped observations into adolescence (10-20 years), early adulthood (21-30 years), and adulthood (31-40 years), and analysed them separately. We identified nuclear families based on shared parentage (with up to two children per nuclear family) and linked them together into extended family units via one of the parents’ twin or sibling. Monozygotic and dizygotic twins in the parent generation were identified via the Norwegian Twin Registry (Nilsen et al., 2012), whereas full siblings were identified through shared parentage as recorded in the population register. For each age group, we identified on average 150,785 extended family units (range: 115,029 – 174,452) that consisted of on average 1,917 (1,719 – 2,228) units linked by monozygotic twins, 2,582 (1,994 – 3,153) units linked by dizygotic twins and 146,286 (109,648 – 170,739) units linked by full siblings. There were on average 1,372,953 (1,029,721 – 1,635,877) observations per age group. See the Supplementary Methods for more details.

### Ethics

The study was approved, and participant consent was waived by the Regional Committee for Medical and Health Research Ethics South-East Norway (REK, approval 2018/434).

### Measures

#### Diagnosis of Psychiatric Disorders

We use psychiatric diagnoses as registered in primary care. Primary care records cover a larger proportion of cases than specialist health care registries and are validated against diagnostic interviews (Torvik et al., 2018). Health records were obtained from the Norwegian Control and Payment of Health Refunds Database (KUHR), which indexes visits to general practitioners. Norwegian health care is heavily subsidized. To receive reimbursement, general practitioners send billing information to the Norwegian Health Economics Administration, along with a diagnosis or reason for the visit coded according to the International Classification of Primary Care (ICPC-2) (WONCA, 2005). Due to this financial scheme, it is unlikely that visits to the general practitioner go unreported. Another study found that >99% of the population had at least one visit to a general practitioner in the study period (Caspi et al., 2024). For our descriptive analyses, we primarily looked at whether offspring had *any* psychiatric disorder, indicated by at least one visit that year registered with a diagnostic code (P70 – P99) in the psychological chapter of the ICPC-2. We also present descriptives for each code separately. See Supplementary Table S3 for the full list of codes and diagnoses. To ensure adequate statistical power for the kinship-based models, we investigated 5-year prevalence instead of 12-month prevalence. This will result in a higher prevalence and therefore more precise estimates. For individuals who were observed for more than five years within the age group, we randomly selected five years and marked them as diagnosed if they had a registered diagnosis in any of the chosen year.

#### Parental Income

Pensionable income records from 1967 to 2017 were provided by Statistics Norway and include salaries and some social benefits. We transformed raw numbers into income ranks based on sex, birth year and tax year. Note that the ranks reflect the entire population and not the subset with children. To account for some of the random year-to-year fluctuations, we calculated the median income rank of each parent between offspring ages 10 to 18. We included the income of parents even if they were not observed for the entirety of this period (e.g., emigrated or deceased). For our descriptive analyses, we used income quartiles based on the distribution of ranks within each offspring birth year.

Measuring income in relation to offspring age rather than parental age meant that parental income rank would differ between offspring of the same parents. We incorporated this into the kinship-based model by modelling a variable parental phenotype with one observation per parent per child.

Because the power of the model relies on the number of monozygotic twin families, which were in short supply, we included monozygotic twin uncles and aunts who themselves did not have children in the respective age group. As we could not measure their income in relation to their offspring’s age, we instead used their median income rank between the age of 30 to 40 (or the median rank between 2007 to 2017 for individuals who had not yet reached the age of 40).

### Statistical Analysis

We estimated prevalences and prevalence ratios by log-binomial regressions for each age with parental income quartile as the predictor and whether the offspring had been registered with any of the relevant codes that year as the outcome variable (1 = diagnosed, 0 = not diagnosed). We did this separately for maternal and paternal income and stratified on offspring sex in follow-up analyses (Supplementary Fig. S1 and S2). We then estimated prevalences for each code separately, also stratified by sex and parental income.

Before modelling the parent-offspring associations, we calculated correlations between various family members (income rank in the parent generation and any psychiatric disorder in the offspring generation) by applying a structural equation model to the data where equivalent relations, such as the parent-offspring relations in the two connected nuclear families, were constrained to be equal. We used a liability threshold model for the offspring phenotype, meaning the correlations in the offspring generation are polychoric correlations, the intergenerational correlations are polyserial correlations, and the correlations in the parent generation are Pearson correlations.

Psychiatric disorders tend to co-occur, and how to categorize different psychiatric conditions is not yet well understood (Kessler et al., 2005; Lahey et al., 2021). Because of these ontological limitations, as well as issues concerning lack of statistical power, we only apply the kinship-based model to the association between parental income rank and any offspring psychiatric disorder.

#### The Kinship-Based Models

The kinship-based model used here (Fig. 1 and Supplementary Fig. S1) is a variant of the iAM-COTS model described in Sunde, Eilertsen & Torvik (Sunde et al., 2024). The iAM-COTS model is a Children-of-Twins model (see also Mcadams et al., 2018; Silberg et al., 2012; Torvik et al., 2020), which is a family of genetically informative structural equation models that extend the classical twin design (Knopik et al., 2017). By including information on the spouses of twins and siblings, it can model and account for indirect assortative mating, and by including information on their children, it can model intergenerational transmission. In this paper, we have adapted the model to handle parental phenotypes that varies over time (cf. Hannigan et al., 2018).

**Figure 1:**
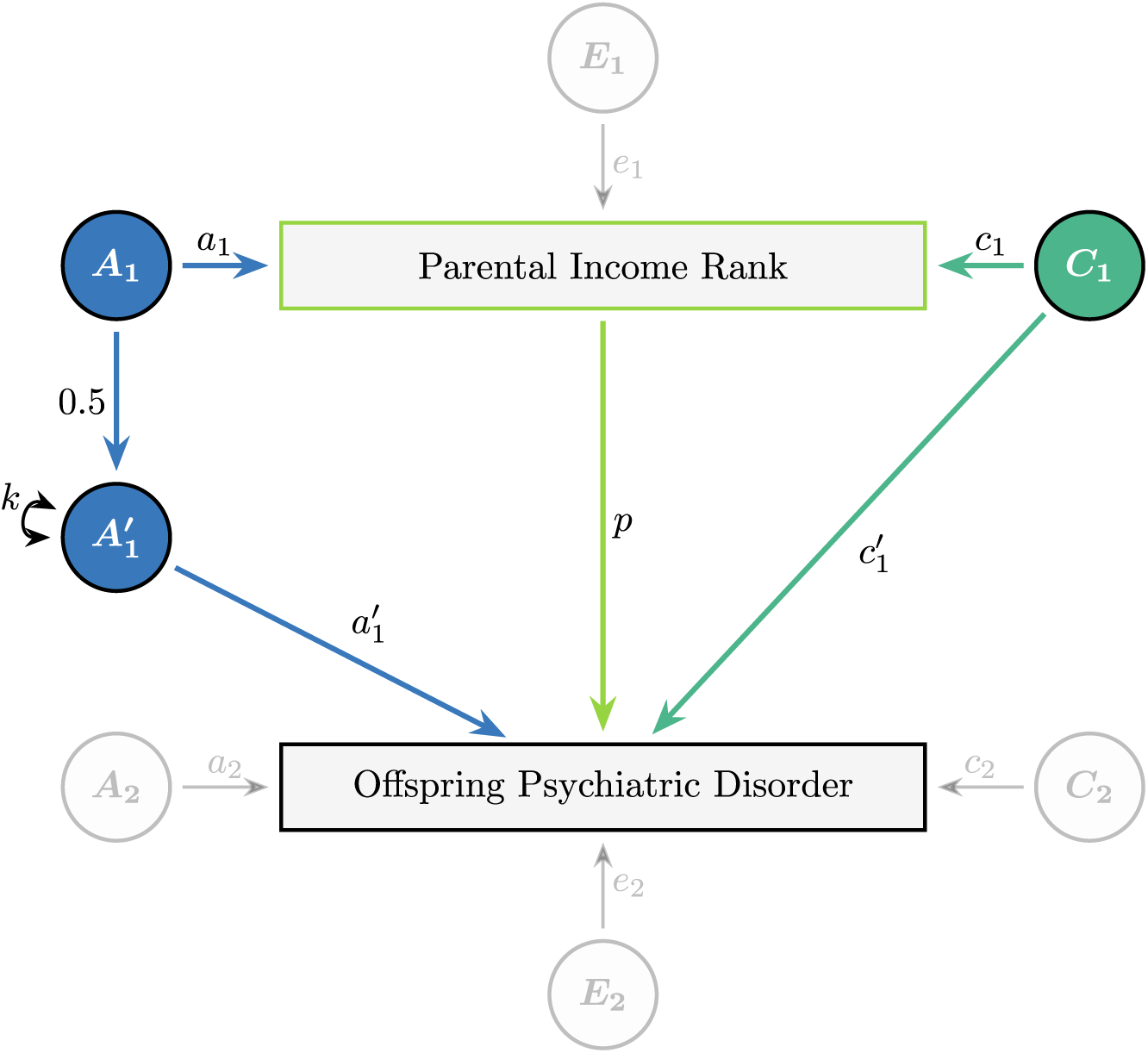
A simplified version of the kinship-based model, showing the relationship between a parental and offspring phenotype (omitting the effects of co-parents and assortative mating). Rectangles represent observed variables and circles represent latent variables. All latent variables have unit variances. The parent-offspring correlation is thought to result from direct phenotypic transmission (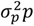, green line), passive genetic transmission 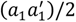, blue line), and passive environmental transmission (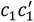, turquoise line). *A* = Additive Genetic Effects, *C* = Shared Environmental Effects, *E* = Non-Shared Environmental Effects. The model is identified by including multiple children and multiple nuclear families linked by monozygotic twins and full siblings. The full biometric model is presented in Supplementary Fig. S3. See Supplementary Table S5 for a complete overview of the parameters in the model and Supplementary Table S6 for the derived equations for the different covariances.

The model decomposes the parent-offspring correlation into three components: *phenotypic transmission*, where paternal income rank is assumed to directly influence offspring risk of psychiatric disorders; *passive genetic transmission*, where offspring resemble their parents due to genetic relatedness; and *passive environmental transmission*, where offspring resemble their parents because they are exposed to the similar environments (assuming these are shared equally in the extended family). The first pathway would be consistent with social causation while the other two pathways would be consistent with social selection. The model can differentiate these pathways because they will have different effects on the avuncular covariance across family types. By comparing the covariances across these different dyads, the model can decompose the parent-offspring covariance into genetic and environmental pathways. The model is described in more detail in the Supplementary Methods, as well as in Sunde et al. (2024). As sensitivity analyses, we also estimated *twin-fixed* effects models (comparing first cousins related through monozygotic twins) and *family-fixed* effects models (comparing siblings within the same nuclear family). These are described in the Supplementary Methods.

#### Software and code

All analyses were conducted in *R 4.2.1* (R Core Team, 2022), using base R functions in conjunction with *data.table* (Barrett et al., 2024), *tidyverse* (Wickham et al., 2019), *flextable* (Gohel & Skintzos, 2022), and *OpenMx 2.20.6* (Neale et al., 2016). The fixed-effects models were estimated using the *clogit* function in the *survival* package (Therneau, 2024). The summary statistics and reproducible code for all figures and tables are available at osf.io/wq8fu/. The supplementary information is also available at osf.io/wq8fu/.

## Results

### Prevalence of psychiatric disorders by age and parental income quartile

The 12-month prevalence of psychiatric disorders increased steadily from age 10 and stabilized around 9.5% in the late twenties (Supplementary Fig. S2). Boys were more likely than girls to have any psychiatric disorder up to the age of 15, after which girls/women were more likely to have any psychiatric disorder. Men had higher prevalence rates of schizophrenia, mental retardation, hyperkinetic disorder, and unspecified disorders, whereas the remaining disorders were more common among women (see interactive figure at https://hfsu.shinyapps.io/prevalence_by_age/, or data files at osf.io/wq8fu/).

Lower parental income was associated with higher prevalence of psychiatric disorders at all ages, although the association decreased somewhat with age (Fig. 2). Comparing the top with the bottom paternal income quartile, the prevalence ratio was 0.47 (95% CIs: 0.45, 0.50) among 10- year-olds, 0.55 (0.54, 0.56) among 20-year-olds, 0.66 (0.64, 0.67) among 30-year-olds, and 0.72 (0.71, 0.74) among 40-year-olds. The absolute prevalence difference was 3.6 percentage points for 10-year-olds, 4.8 points for 20-year-olds, 4.1 points for 30-year-olds, and 3.0 points for 40-year-olds. Results were similar for maternal income, but with smaller differences among the oldest age groups.

**Figure 2:**
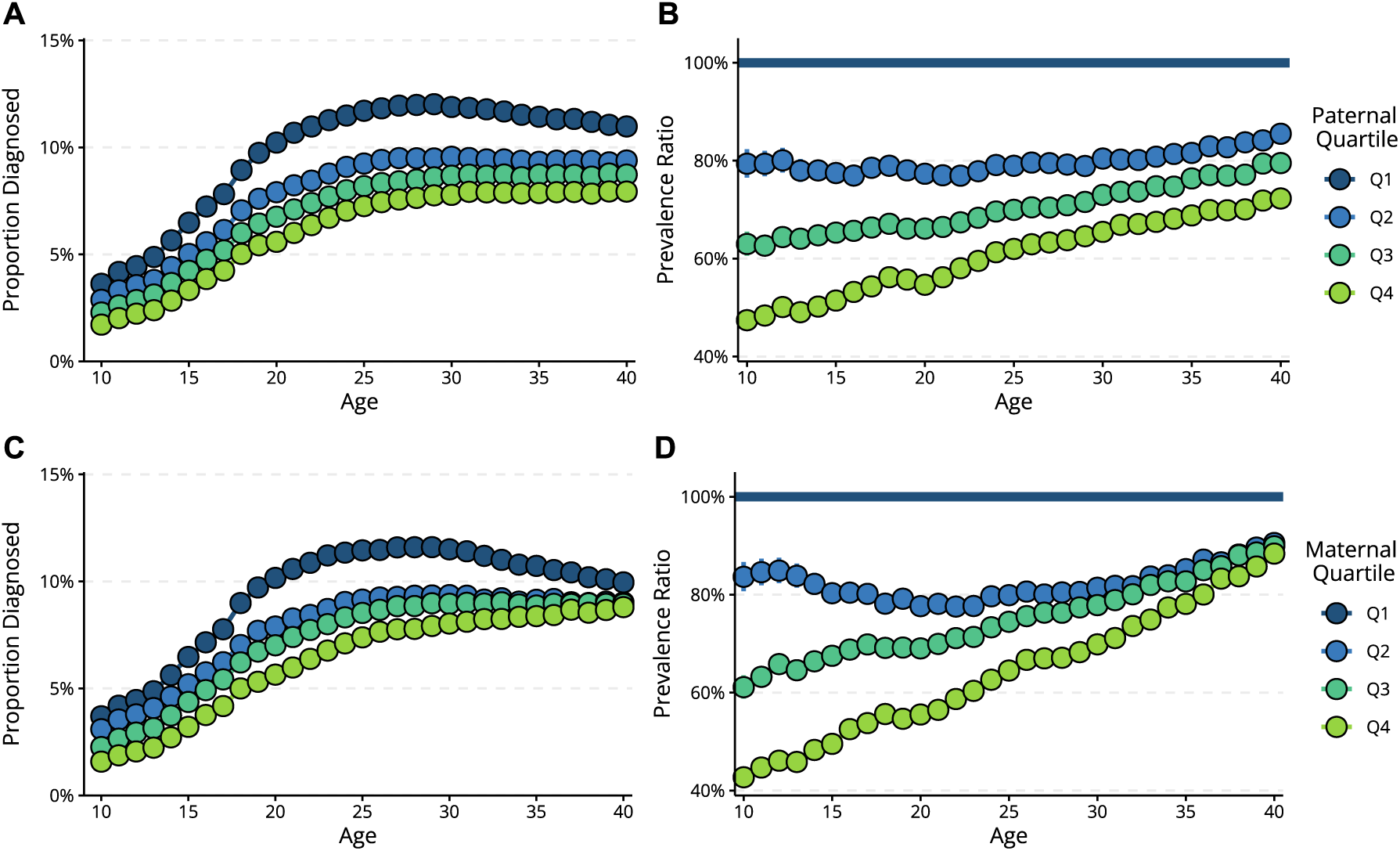
Psychiatric disorders across age and parental income quartile. 95% CIs are too small to be visible in this figure. The top panels present results for paternal income quartile (**A**: prevalences, **B**: prevalence ratios), and the bottom panels present results for maternal income quartile (**C**: prevalences, **D**: prevalence ratios).

There were large sex differences in prevalence and developmental trajectories of psychiatric disorders, but despite this, the associations with parental income rank were similar for both men and women throughout the age range (Supplementary Fig. S2 – S3). Likewise, prevalence and developmental trajectories differed between psychiatric disorders, but almost all psychiatric disorders were associated with parental income (Fig 3). Eating disorders stood out as the only disorder without a clear parental income gradient.

**Figure 3:**
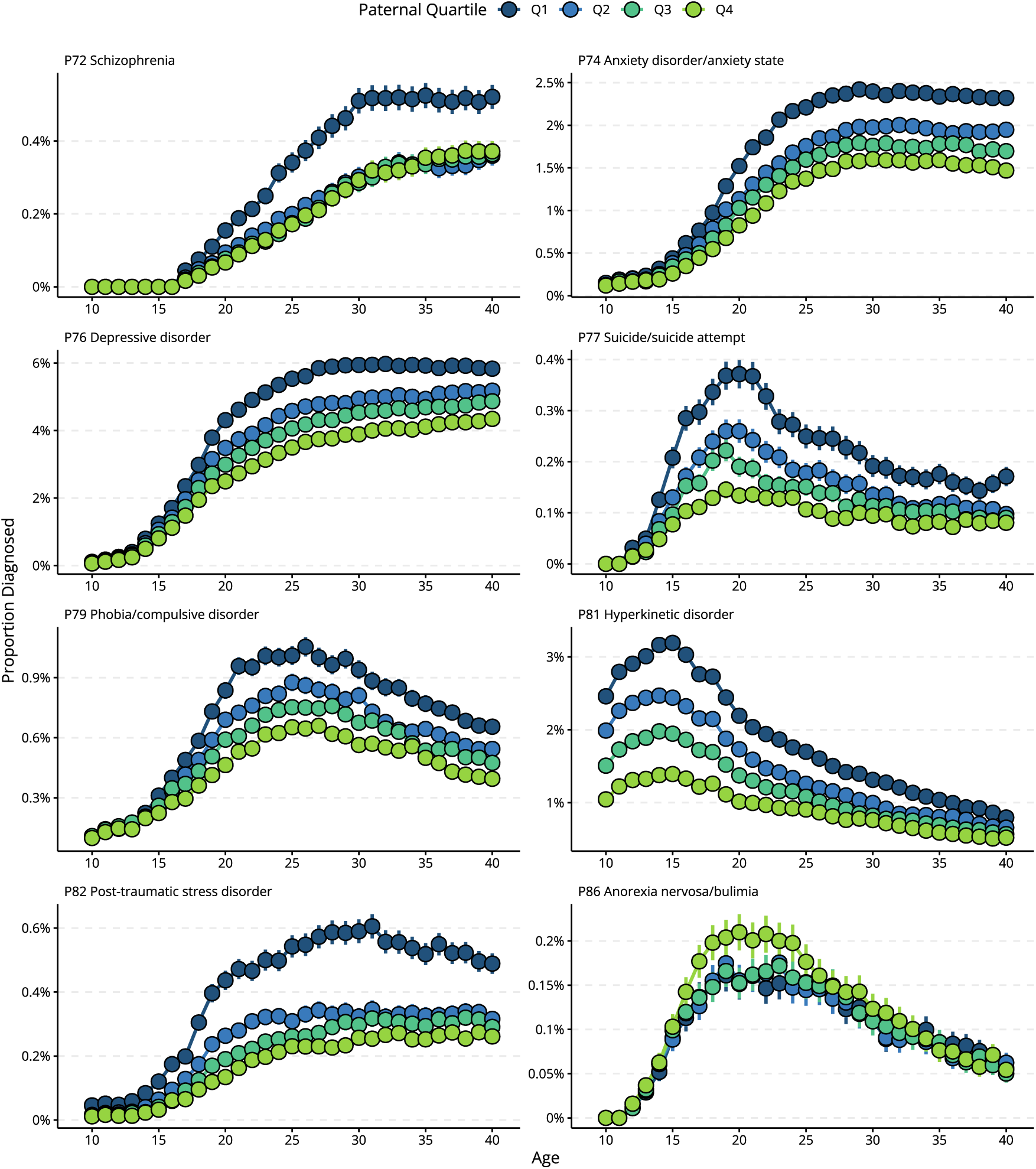
Proportion diagnosed (incl. 95% CIs) with a selection of specific psychiatric disorders the year they turn a given age, stratified by paternal income quartile. Note that the scaling of the y-axes varies so that differences in low-prevalence disorders become visible. Results for all investigated diagnostic codes, including stratification by maternal income quartile and by offspring sex, are presented in an interactive figure at https://hfsu.shinyapps.io/prevalence_by_age/, and in data files at osf.io/wq8fu/.

### Intergenerational correlations across family type

In Fig. 4, we present polyserial correlations between parental income rank and any psychiatric disorder for parent-offspring pairs and avuncular pairs (offspring and their parent’s sibling), across age groups and family types (see Supplementary Table S4 for correlations among other family members). The parent-offspring correlation decreased with age. Among families in the full sibling group (the group with the highest sample size and therefore smallest confidence intervals), the parent-offspring correlation was −.146 (95% CI: −.150, −.143) in adolescence, −.100 (−.100, −.093) in early adulthood, and −.060 (−.064, −.056) in later adulthood. Parent-offspring correlations were similar in other family types, albeit with larger confidence intervals. The avuncular correlation was generally smaller, but this dependent on age group and family type. In full sibling families, the avuncular correlation was about half the parent-offspring correlation across age groups, which is expected if the parent-offspring correlation could be fully attributed to genetic similarity (although combinations of different forms of environmental transmission can also give similar expectations). In monozygotic twin families, where offspring are equally genetically related to their uncle/aunt as to their parent, the avuncular correlation was smaller than the parent-offspring correlation among adolescents, while but about equal among adults (albeit with wide confidence intervals).

**Figure 4:**
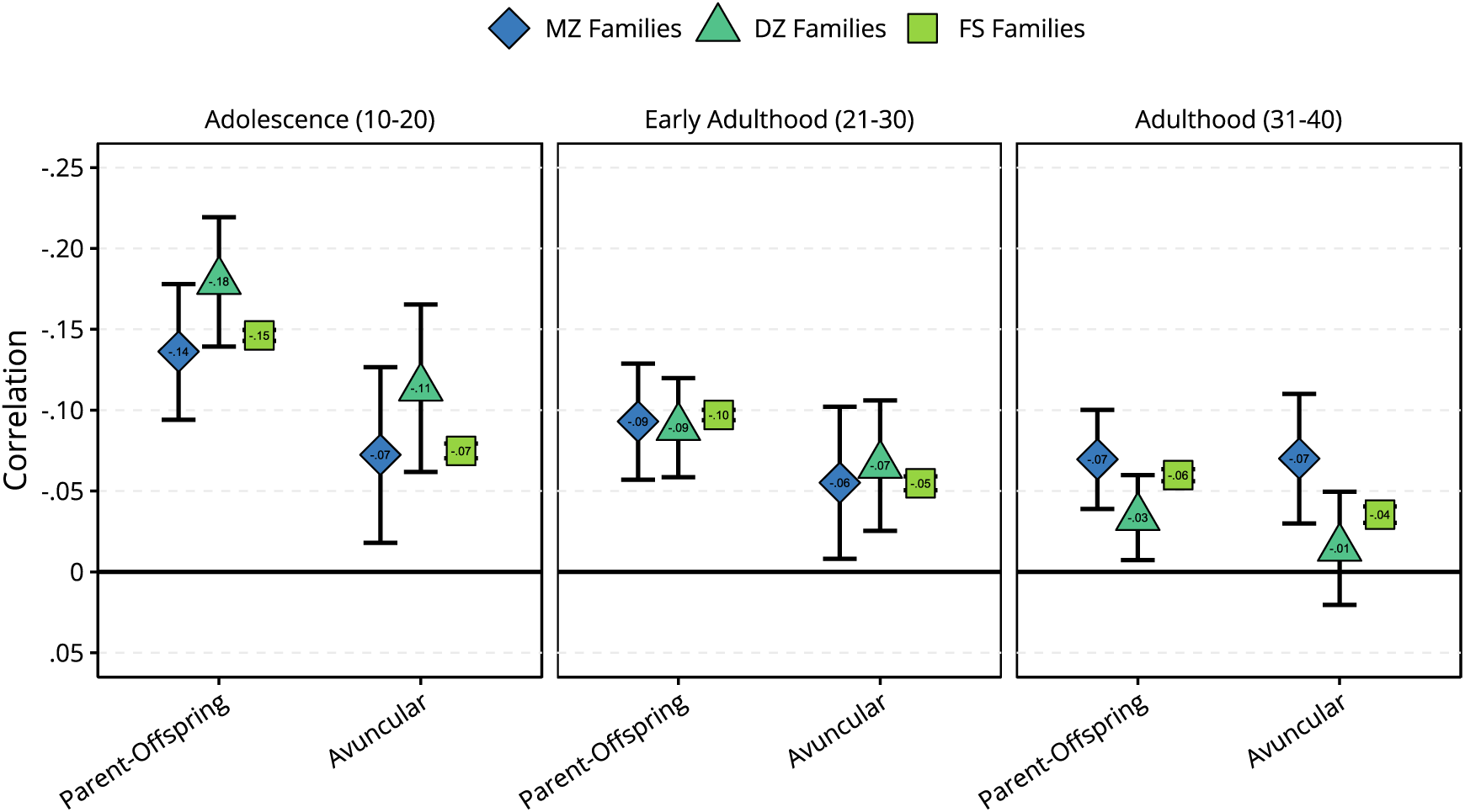
Polyserial correlations between parental or avuncular (uncle or aunt) income rank and offspring psychiatric disorder, stratified by age group and family type (i.e., whether uncle/aunt is a monozygotic twin (MZ), dizygotic twin (DZ), or a full sibling (FS) of the parent). The y-axis is reversed to make interpretation more intuitive. The numbers behind this figure are listed in Supplementary Table S4.

### The Kinship-Based Models

The results of the decomposition of the parent-offspring correlation are presented in Fig. 5. For all three age groups, we found significant contributions from both passive environmental transmission (blue) and especially passive genetic transmission (turquoise). We found evidence of phenotypic transmission (green) from parental income rank among adolescents, where it accounted for 38.6% (95% CI: 29.0%, 48.2%, *p* < .001) of the parent-offspring correlation (with 47.7% (35.7%, 59.7%) being ascribed to passive genetic transmission and 13.7% (5.9%, 21.5%) being ascribed to passive environmental transmission). Phenotypic transmission was not statistically significant in the two other age groups (early adulthood: Δ-2LL = 1.79, Δdf = 1, *p* = .181, adulthood: Δ-2LL = 0.11, Δdf = 1, *p* = .736). Models constrained to assume *direct* assortative mating yielded different results (Supplementary Fig. S4), but this resulted in significantly worse fit (e.g., adolescence, Δ-2LL =4293.99, Δdf = 4, *p* < .001, see also Supplementary Tables S11-S13). All estimated and derived parameters are presented in Supplementary Tables S7 to S10, whereas model comparison statistics are presented in Supplementary Tables S11 to S13.

**Figure 5:**
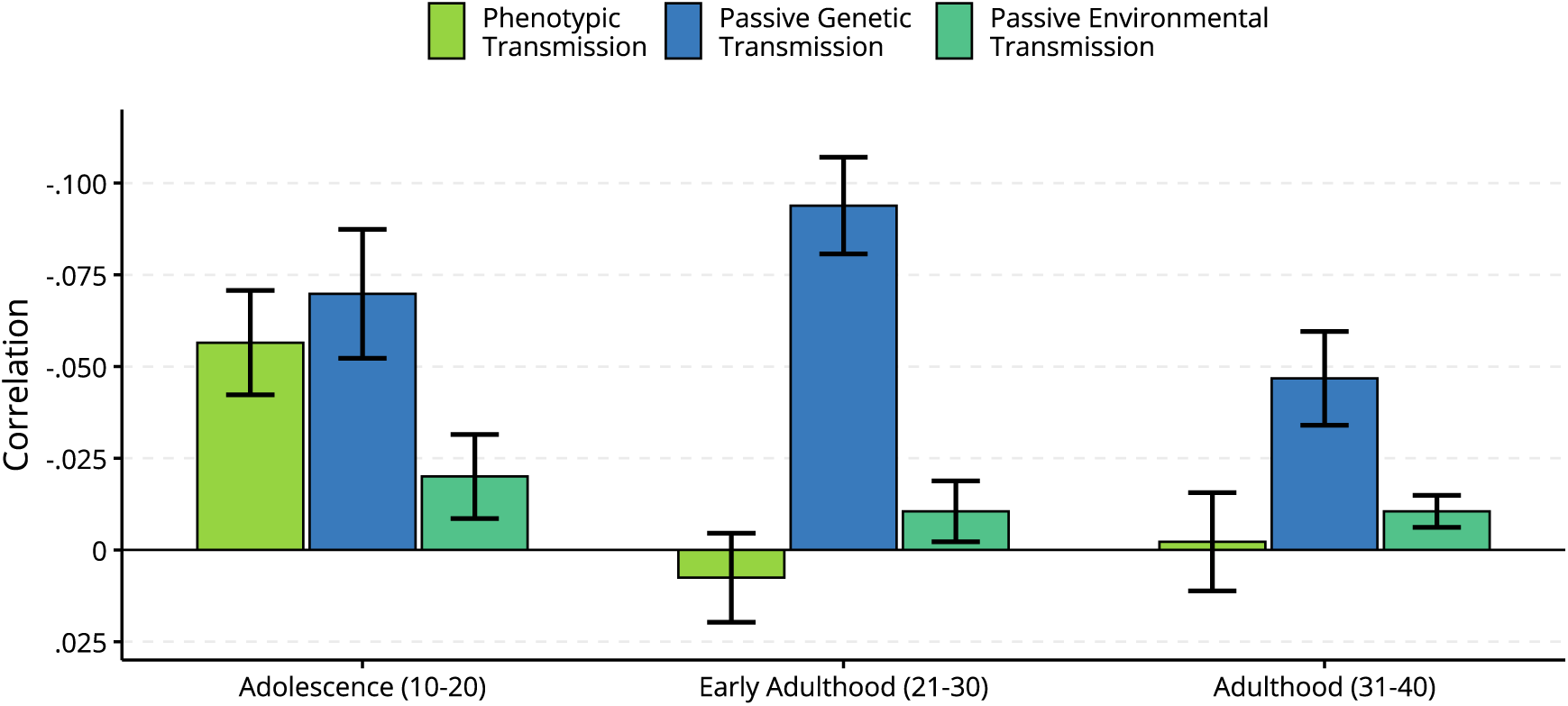
The decomposition (incl. 95% CIs) of the correlation between parental income rank and any offspring psychiatric disorders across offspring age groups. The correlations are assumed to reflect phenotypic transmission (i.e., direct effects of parental income rank, green), passive genetic transmission (blue), and passive environmental transmission (turquoise). The total correlation is the sum of each component. The y-axis is reversed to make interpretation easier, so that a taller column means a larger negative correlation (lower income rank → higher risk of psychiatric disorders). Results are also presented in Supplementary Table S9.

The heritability of having any psychiatric disorders was estimated to 52% (95% CIs: 46%, 59%) among adolescents, 49% (47%, 52%) among adults aged 21 to 30, and 40% (38%, 42%) among adults aged 31 to 40. The genetic correlation between income and psychiatric disorders were estimated to be moderate, ranging from −.24 (−.29, −.19) among adolescents to −.37 (−.42, −.33) among adults aged 21 to 30.

### Fixed-effects models

Results from the fixed-effects models are presented in the Supplementary Results. Among adolescents, we found significant family-fixed effects (OR = 0.964, 95% CI: 0.943, 0.985, *p* < .001), but not twin-fixed effects (OR = 0.907, 95% CI: 0.924, 1.003, *p* = 0.57). In the two other age groups (early adulthood and adulthood), neither twin-fixed effects nor the family-fixed effects were statistically significant (Supplementary Fig. S5, Supplementary Table S14).

## Discussion

We have described how parental income rank was associated with offspring risk of psychiatric disorders across 30 years from early adolescence and used kinship-based models to differentiate between social causation and social selection. We found that lower parental income was associated with higher prevalence of psychiatric disorders at all ages from age 10 to 40, with a moderate decrease with age. Although the prevalence and developmental trajectory varied between sexes and between different disorders, lower parental income was associated with higher prevalence of nearly all psychiatric disorders for both men and women. The kinship-based models indicated that social selection played a large role in explaining the parent-offspring correlation, through significant effects of both passive genetic transmission and passive environmental transmission. However, we found evidence that indicated social causation during adolescence, suggesting a transient effect of parental income rank (or a within-family correlate) on offspring psychiatric disorders.

While no practically applicable method exists that can decide the causation-selection issue once and for all, our finding corroborates several lines of evidence that hint at a causal effect of parental income – or a correlate of income – on adolescent psychiatric disorders (Akee et al., 2018; Costello et al., 2003; Kinge et al., 2021). This triangulation of methods with different sets of strengths and weaknesses makes us confident that social selection alone cannot explain the entirety of this intergenerational association. Furthermore, the consequence of a causal effect depends on the variance of the putative cause. Because Norway has relatively small income disparities (OECD, 2022; The World Bank, 2022), the variance in parental income is lower than in many other countries. As a result, the proportion of variance in psychiatric disorders explained by direct effects of parental income may also be smaller. If a causal relationship is observed in this context, it is plausibly more pronounced in societies with greater inequality and more barriers to healthcare access. Future studies may want to investigate how our findings generalize to other cultural contexts.

### On effects across different age groups

A key aim of this study was to investigate whether direct effects of parental income, if any, changed with offspring age, as this would plausibly explain some of the conflicting findings from the literature. Our findings are consistent with this explanation, adding to numerous lines of evidence suggesting that phenotypic stability over time is often due to genetic factors, whereas environmental effects tend to vary from age to age (Plomin et al., 2016). Phenotypic transmission, evident among adolescent offspring, was not found among adult offspring. Direct effects of parental income may therefore be transient. That is, lower parental income rank (or a correlate of income) could increase the risk of psychiatric disorders when the children still live with their parents, but this increased risk does not seem to follow children after they have moved out into different environments. This could explain why Kinge et al. (2021), who investigated children, and Sariaslan et al. (2021), who investigated adults, reached different conclusions, although alternative explanations such as methodological differences are also plausible (Biele et al., 2022). Alternatively, the lack of effects that last into adulthood can be a false negative attributed to, for example, non-linear effects or other violated assumptions, or because social causation is only detectable with larger variation in economic resources than seen in Nordic cultural contexts.

### On Passive Genetic Transmission

Regardless of whether effects truly persist into adulthood, social selection is clearly an important contributor to the parent-offspring correlation. The kinship-based models indicated that passive genetic transmission accounted for a large proportion of this. That is, there was an estimated genetic correlation ranging from −.24 to −.37 between genetic variants that were associated with income rank and variants that were associated with psychiatric disorders common in that age group. Similar genetic correlations between income and psychiatric disorders have also been found in other studies (Abdellaoui & Verweij, 2021; Kweon et al., 2025; Marees et al., 2021). Because offspring are genetically related to their parents, parental income rank and offspring psychiatric disorders become phenotypically correlated in family members. The cause of this genetic correlation remains unknown, but a plausible explanation is that propensity for psychiatric disorders runs in families (Polderman et al., 2015) and psychiatric disorders influence educational achievement (Nordmo et al., 2022; Sunde et al., 2022) and consequently salaries (Markussen et al., 2020; Miller, 1998). Other plausible mechanisms include cognitive and personality traits, which may influence socioeconomic status and mental health (Gupta et al., 2024; Nordmo et al., 2024). Genetic correlations between socioeconomic indicators and psychiatric conditions appear to play a key role in explaining psychiatric comorbidity (Marees et al., 2021). Other causal processes, such as assortative mating across socioeconomic and psychiatric traits (Border et al., 2022), are also plausible. Future research should attempt to disentangle the various mechanisms that could lead to this genetic correlation.

### On Passive Environmental Transmission

Unlike previous studies, our analysis differentiates genetic and environmental sources of intergenerational confounding. We find that passive environmental transmission was a significant contributor across all age groups. These are income-associated environmental factors shared by parental siblings and twins which are also associated with risk of psychiatric disorders among their offspring. The specific environmental factors involved remain unclear, but could involve neighbourhood quality, values related to culture, religion, or social class, and exposure to adversity, to the degree these are shared by extended family members. Future research is needed to identify which shared environmental influences drive these associations. Regardless, these findings highlight that it is important to account for multiple sources of confounding in intergenerational studies.

### Differences between register-based and interview-based prevalences

Prevalence rates based on medical records stored in primary care health registers are generally lower than those reported in interview studies. For example, whereas Kessler et al. (2005) reported the 12-month prevalence of any disorder at 26.2%, we find a 12-month prevalence at around 9.5%. A Norwegian study found high genetic correlations (median *r* = .81) between health registries and diagnostic interviews, suggesting they are the result of similar etiological influences (Torvik et al., 2018). They therefore attributed the difference in prevalence between interviews and health registries as a difference in threshold, not a difference in phenotype. The different prevalences found in health records compared to diagnostic interviews are therefore unlikely to lead to different conclusions for the role of parental income. This also suggest that diagnostic interviews may have a threshold that often captures cases that are not severe enough to seek medical attention.

### Considerations when interpreting the kinship-based model

The kinship-based model estimates various pathways assumed to reflect causal effects, but like any structural equation model, the results cannot be interpreted causally without caveats. Phenotypic transmission, labelled *p* in Fig. 1, captures aspects of the parent-offspring correlation that cannot be attributed to genetic relatedness or extended family environments (Mcadams et al., 2018). This includes direct effects of parental income rank on offspring psychiatric disorders (which is what the model assumes), but plausibly also effects of offspring psychiatric disorders on their parents’ income rank. While temporal precedence makes the latter somewhat unlikely in our study, at least for the later age groups, it cannot be ruled out. Furthermore, the model does not allow us to conclude that effects are directly attributable to the *observed* phenotype. Confounding factors that are not captured by the genetic and extended family effects will look like phenotypic transmission. This includes other parental traits that influence their income rank, and which also influence their offspring’s risk of psychiatric disorders. As such, our finding of significant phenotypic transmission during adolescence can only be interpreted as evidence that *suggests* that income rank *or a correlate* likely influences offspring psychiatric disorders (while acknowledging that reverse causation or more obscure forms of confounding are also technically possible).

The kinship-based model relies on many of the same assumptions as most twin models, such as no gene-environment interactions, no gene-environment correlations in the parent generation, and the equal environments assumption. In a children-of-twins model like the one used here, the equal environments assumption is further expanded to include that offspring of monozygotic twins are not exposed to relevant environmental factors shared by extended family members more than offspring of dizygotic twins and full siblings (Mcadams et al., 2018). Gene-environment correlations are assumed absent in the parent-generation, but in the offspring generation, it follows from co-occurring genetic and phenotypic transmission (accounting for 0 to 1.8% of the variance). Finally, the kinship models estimate population-average effects. Any non-linear or localized effects will go unnoticed unless they are large enough to influence the population average effect. While it is possible to incorporate interactions in children-of-twins models (Badini et al., 2024), it remains unclear if this is feasible when also modelling indirect assortative mating and dichotomous variables. Future research should investigate the lowest income brackets and poverty, which may have qualitatively different causal effects than general variation in income rank.

### Limitations and outstanding questions

Despite using a genetically informative design and primary health care data on an entire population, our study has some limitations, and some questions remain unanswered. First, our measure of psychiatric disorders was dichotomous, which precludes investigating any sub-threshold effects (e.g., symptoms) and reduces power. Second, the statistical power in the kinship-based models is limited by the number of monozygotic twins and the dichotomous health register data. Because specific disorders necessarily have a lower prevalence than the sum of any psychiatric disorder, the estimated correlations with parental income are less accurate and consequently more difficult to decompose. When we attempted to run the model on specific disorders, it would often fail to converge and provided unreliable results. We therefore only decomposed the association between parental income rank and any psychiatric disorder, although we recognize that the specific decompositions likely vary across different psychiatric disorders. The purported age-effects in this study may be attributable to different disorders being common in different age groups. On the other hand, we found similar results in the fixed-effects models when only looking at internalizing disorders (see Supplementary Results). Third, individual differences in help-seeking behaviour may influence the likelihood of being registered with a diagnosis, especially for cases near the diagnostic threshold. To the extent that this is associated with parental income rank, this will confound the results. Fourth, the twin registry is based on informed consent and may therefore suffer from some participation bias. Fifth, the model assumes that the sources of variation in the parental phenotype are the same for mothers and fathers, and that they have an identical effect on the offspring. There was not enough power to relax these assumptions. Future research may want to undertake a more thorough investigation into sex differences in the sources of income rank variation and how they differentially affect offspring. Furthermore, more research is needed into how different operationalizations of parental income and socioeconomic status relate to psychiatric disorders, and what related factors, if any, are decisive in this association.

## Conclusion

To conclude, we found a clear association between parental income rank and prevalence of offspring psychiatric disorders that persisted into adulthood. The association is present for both men and women and for most psychiatric disorders. By using genetically informative structural equation models, we found results that suggested that a large part of the association was due to passive forms of transmission, mainly genetic transmission (i.e., the *social selection* hypothesis). This would mean that offspring of parents with lower income rank are at an increased risk of psychiatric disorders in the current cultural context partly because the set of traits that result in lower income rank are inherited and overlaps with the set of traits that influence risk of psychiatric disorders. Future research should attempt to identify why these traits overlap or co-occur. After controlling for shared genetic and environmental factors, we found evidence of phenotypic transmission during adolescence but not adulthood. Hence, we found results indicative of transient effects of parental income rank (or a correlate of income rank) on offspring psychiatric disorders, supporting the *social causation* hypothesis among adolescents, but not adults.

## Acknowledgements

This work was supported by the Research Council of Norway (grants number 273659 and 300668). This work was partly supported by the Research Council of Norway through its Centres of Excellence funding scheme (grant number 262700). Data on twin zygosity were obtained from the Norwegian Twin Registry, Norwegian Institute of Public Health. The funders of this study had no role in study design, data collection, data analysis, data interpretation, or writing of the report.

## Data Availability

The data for this study encompasses tax records, health records, and demographic information for the entire Norwegian population. Researchers can access the data by application to the Regional Committees for Medical and Health Research Ethics and the data owners. The authors cannot share these data with other researchers.

## Author Contributions

Conceptualisation: H.F.S. and F.A.T.; Methodology: H.F.S., E.M.E. and F.A.T. Formal analysis: H.F.S, with supervision from E.M.E. and F.A.T.; Data Curation: H.F.S.; Writing – Original Draft: H.F.S.; Writing – Review and Editing: All authors; Visualization: H.F.S. and E.M.E.; Supervision: F.A.T. and E.M.E.; Project administration and funding acquisition: F.A.T.; All authors provided critical feedback, discussed the results, helped shape the manuscript, and approved the final paper for submission. H.F.S. had full access to the data and takes responsibility for the work.

