## Supplementary Information for "Parental income and psychiatric disorders from age 10 to 40: a genetically informative population study"

### Table of Contents

Notes on the supplementary materials: 3

Supplementary Methods 4
Supplementary Results 7

Figure S1: Path diagram for the kinship-based model 8
Figure S2: Prevalence of any psychiatric disorder by sex and income quartile 9
Figure S3: Prevalence ratios for any psychiatric disorder by sex and parental income 10
Figure S4: Decomposition of parent-offspring correlation when assuming direct assortment 11
Figure S5: Fixed-effects coefficients (any psychiatric disorder) 12
Figure S6: Fixed-effects coefficients (Internalizing and ADHD) 13

Table S1: Number of observations per age 14
Table S2: Number of observations per age group (kinship models) 15
Table S3: Psychological diagnostic codes in ICPC-2 16
Table S4: Correlations in extended family 18
Table S5: Parameters in the kinship-based model 20
Table S6: Equations implied by the iAM-COTS model in Figure S3 21
Table S7: Estimated Free Parameters in the Kinship-Based Model 23
Table S8: Standardized Variance Components 24
Table S9A: Decomposition of Parent-Offspring Correlation 25
Table S9B: Decomposition of Parent-Offspring Correlation (percentages) 26
Table S10: Other derived parameters 27
Table S11: Model Comparisons for Adolescence (10-20) 28
Table S12: Model Comparisons for Early Adulthood (21-30) 29
Table S13: Model Comparisons for Adulthood (31-40) 30
Table S14 Fixed-Effects Models (Any Psychiatric Disorder) 31
Table S15 Fixed-Effects Models (Internalizing Conditions) 32
Table S16 Fixed-Effects Models (ADHD) 33

#### Notes on the supplementary materials:

Every figure and table (except the path diagrams) are made in R using *ggplot2* and *flextable*. The code to reproduce each figure and table, along with the relevant summary statistics, are available at osf.io/wq8fu/.

Prevalences where the number of cases in any of the sub-categories are below 20 cases are replaced with 0 in the summary statistics because of privacy concerns related to rare cases.

Prevalences for every diagnostic code across age, sex, and paternal/maternal income quartiles are available in an interactive figure at https://hfsu.shinyapps.io/prevalence_by_age/, or as data files at osf.io/wq8fu/.

#### Supplementary Methods

##### Constructing the extended family units

For the kinship-based models, we only used data on individuals who were observed for at least 5 years during the relevant ages (i.e., alive and living in Norway during adolescence, early adulthood, and/or adulthood). For each age group, we identified nuclear families (i.e., sets of parents and their children) with children in the respective age groups. For nuclear families with more than two eligible children, we randomly selected two children. This randomization procedure also randomized the order of children, so that first-borns were not always considered the first child (to avoid complications from parity effects interfering with locations in the covariance matrix). We then paired nuclear families into extended family units by linking together same-sex twins and full siblings in the parental generation. Monozygotic and dizygotic twins were identified via the Norwegian Twin Registry (Nilsen et al., 2012), whereas full siblings were identified through shared parentage as recorded in the population register. Twins with unknown zygosity (identified via shared parentage and birth month) were not considered eligible pairs in the full sibling group. We first linked together all nuclear families that were linked via monozygotic twins, then via dizygotic twins, before joining the remaining nuclear families via a random same-sex full sibling. Nuclear families that formed part of extended family units on both the maternal and paternal side were randomly allocated to one of them. For each age group, we identified on average 150,785 extended family units (range: 115,029 – 174,452) that consisted of on average 1,917 (1,719 – 2,228) units linked by monozygotic twins, 2,582 (1,994 – 3,153) units linked by dizygotic twins and 146,286 (109,648 – 170,739) units linked by full siblings. There were on average 1,372,953 (1,029,721 – 1,635,877) observations per age group. See Supplementary Table S2 for detailed sample sizes for each age group.

##### The iAM-COTS model.

The model we used here is the iAM-COTS model (Sunde et al., 2024), adapted to incorporate variable parental phenotypes (Supplementary Figure S1). The model takes as its input 12 variables reflecting the observed phenotypes of a pair of twins/siblings, their spouses, and their children (up to two per nuclear family). There is one observed offspring phenotype per child, and one observed parental phenotype per parent per child (two children → two observed phenotypes for each parent). The 12 variables yield a 12x12 covariance matrix which differs between family types (i.e., monozygotic twins versus dizygotic twins / full siblings). The goal of the model is to reduce these matrices, which normally contain 78 observed statistics each, down to just 14 free parameters. These parameters represents influences on parents: $a_1$, $c_1$, $e_{1b}$, and $e_{1w}$; similarity between partners and in-laws: $\tilde{a}_1$, $\tilde{c}_1$, $\tilde{e}_1$, and $\mu$; associations between parents and children: $p$, $a_1'$, and $c_1'$; and child-specific influences $a_2$, $c_2$, and $e_2$ (see Supplementary Table S5 for a full description). Because dizygotic twins and full siblings were equally correlated (e.g., adolescence: $r_{DZ}$ = .28 [.24, .32], $r_{FS}$ = .27 [.27, .27]), we combined these groups in the model. It is possible to include a separate twin-shared environmental factor to the model, but this did not significantly improve fit in any of the age groups (all $p$'s > .256, see Supplementary Tables S11-

S13). Because co-siblings-in-law were more correlated than siblings-in-law, which comes about when siblings tend to mate with similar partners beyond their tendency to mate with partners similar to themselves (Eaves, 1979), we added two parameters, $\zeta_{MZ}$ and $\zeta_{FS}$, to account for the excess correlation between co-siblings-in-law (as described in Sunde et al., 2024).

To reduce the observed statistics down to 16 parameters, the cells in the 12x12 covariance matrices are replaced by equations made up of the free parameters, known values (e.g., relatedness), and other derived parameters. The model then uses Maximum Likelihood estimation to find values for the free parameters that, when entered into these equations, produce expected covariance matrices that resemble the observed covariance matrices as closely as possible. When the model produces values that result in expected covariance matrices that resemble the observed covariance matrices, the model is said to be a good fit. The equations are derived using path tracing rules (Balbona et al., 2021; Keller et al., 2009; Sunde et al., 2024; Wright, 1934) following the full path diagram in Supplementary Fig. S3 and are listed in Supplementary Table S6. The key covariance of interest, namely the parent-offspring covariance, can be expressed using the following equation:

$$Cov(Parent, Offspring) = \sigma_p^2 p + \frac{a_1 a'_1}{2} + c_1 c'_1 + \sigma_{PS} \mu \lambda_{SO} \quad (1)$$

Here, $\sigma_p^2$ refers to the variance of the parental phenotype, $\sigma_{PS}$ refers to the within-person covariance between the observed parental phenotype and the sorting factor (representing the traits associated with the phenotype that partners are assorting on), and $\lambda_{SO}$ refers to the covariance between the parental sorting factor and offspring phenotype (Sunde et al., 2024). The equation has four components: The first three components represent the three causes of similarity described above (direct phenotypic transmission, passive genetic transmission, and passive environmental transmission). The fourth component represents the inflation of the other three components that results from assortative mating. In Fig 5, this inflation is included in the respective components (see also Supplementary Table S9).

The offspring phenotype was dichotomous, and therefore modelled with a liability threshold model, meaning we assume the presence or absence of psychiatric disorders reflects an underlying normally distributed liability. We constrained the offspring phenotypic mean ($\overline{X_o}$) to zero and the offspring unique environmental effect ($e_2$) to one, and freely estimated a threshold ($\theta$). The variance components and related paths were standardized afterwards.

The iAM-COTS model estimates and accounts for indirect assortative mating by estimating the degree of genetic homogamy ($\tilde{a}_1$), social homogamy ($\tilde{c}_1$), and idiosyncratic homogamy ($\tilde{e}_1$), and adjusting the intergenerational paths accordingly (Sunde et al., 2024). For example, under genetic homogamy, partners will have correlated genotypes, which increases the similarity between parents and offspring owing to genetic similarity. Many prior models that accounted for partner similarity assumed *direct* assortment, where the degree of genetic, social and idiosyncratic homogamy equals the relative importance of genetic, social and idiosyncratic factors. In an earlier version of this manuscript, we used a model that made this assumption, and got results that differed somewhat from those presented in the main manuscript. In general, passive genetic transmission was estimated to be more important, whereas direct phenotypic transmission was estimated to be less important, and even significant in the opposite direction among older age groups. The earlier version of the manuscript is available at https://osf.io/6324g/. It is possible to reproduce those results and test for significant differences in fit by constraining the model to direct assortment (as described in Sunde et al., 2024). This results in similar results as before (e.g., reversed phenotypic transmission in early adulthood), but with significantly worse fit (see Supplementary Figure S4 and Supplementary Tables 11-S13).

**Sensitivity analyses: Fixed-effects models**

To supplement the kinship-based models, we estimated fixed-effects models in each age group using within-family variation in income rank as the predictor, scaled so that a unit increase corresponds to 10% percentiles higher income rank:

$$\log\left(\frac{P(Y_{ij}=1)}{P(Y_{ij}=0)}\right) = \beta_1 IncomeRank_{ij} + \beta_2 BirthYear_{ij} + \lambda_j \tag{2}$$

Here, $Y_{ij}$ denotes whether individual $i$ in family $j$ has a psychiatric diagnosis (1) or not (0); $\lambda_j$ represents stratum-specific fixed effects, which conditions out characteristics shared by the relevant family members (depending on which stratum is used); and $\beta_1$ represents the effect of within-stratum variation in income rank. All models were adjusted for birth year to account for parity effects (this was accounted for by design in the kinship-based models, see above). The models were estimated as conditional logistic regressions using *clogit* in R's *survival* package.

In the first model (henceforth *twin-fixed effects*), we compared first cousins related through monozygotic twins asking whether offspring of the twin-parent with the higher income rank have a lower risk of psychiatric disorders. In the second model (henceforth *family-fixed effects*), we analysed full siblings within the same nuclear family asking whether the offspring who experience higher parental income rank when they were between ages 10 to 18 have a lower risk of psychiatric disorders. For comparison, we also estimated the population-level association using normal logistic regressions, as well as fixed-effect models comparing first cousins related through full siblings. We used any psychiatric disorder as the primary outcome variable. Additionally, we used ADHD and internalizing disorders as alternative outcome variables. ADHD was defined using any registration with ICPC-2 code P81 (hyperkinetic disorder), whereas internalizing conditions were defined as any registration with anxiety and/or depressive symptoms or disorders (ICPC-2 codes P01 feeling anxious/nervous/tense, P03 feeling depressed, P74 anxiety disorder, and P76 depressive disorder).

### Supplementary Results

#### Sensitivity analyses: fixed-effects models

Results from the fixed-effects models are presented in Supplementary Fig. S5. Among adolescents, the population-level analysis showed that a decile increase in parental income rank was associated with a reduced risk of any psychiatric disorder (OR = 0.902, 95% CI: 0.899, 0.905, $p < .001$). When comparing first cousins related through monozygotic twins, the point estimate was similar albeit with wider confidence intervals (OR = 0.907, 95% CI: 0.924, 1.003). It was nominally non-significant ($p = .057$). When comparing full siblings within the same nuclear family, the sibling with the higher parental income rank were at reduced risk of psychiatric disorders (OR = 0.964, 95% CI: 0.943, 0.985, $p < .001$). In the two other age groups (early adulthood and adulthood), neither twin-fixed effects nor the family-fixed effects were statistically significant (Supplementary Fig. S5, Supplementary Table S14).

Internalizing conditions and ADHD had overall similar results but with wider confidence intervals (Supplementary Fig. S6, Supplementary Tables S15-S16). None of the twin-fixed effects were significant, except for ADHD in early adulthood (OR = 0.813, 95% CI: 0.664, 0.996, $p = .046$). The family-fixed effects were significant for internalizing disorders among adolescents (OR = 0.959, 95% CI: 0.936, 0.982, $p < .001$) and reversed in early adulthood (OR = 1.017, 95% CI: 1.001, 1.033, $p = .037$). All other family-fixed effects were non-significant.

#### Figure S1: Path diagram for the kinship-based model

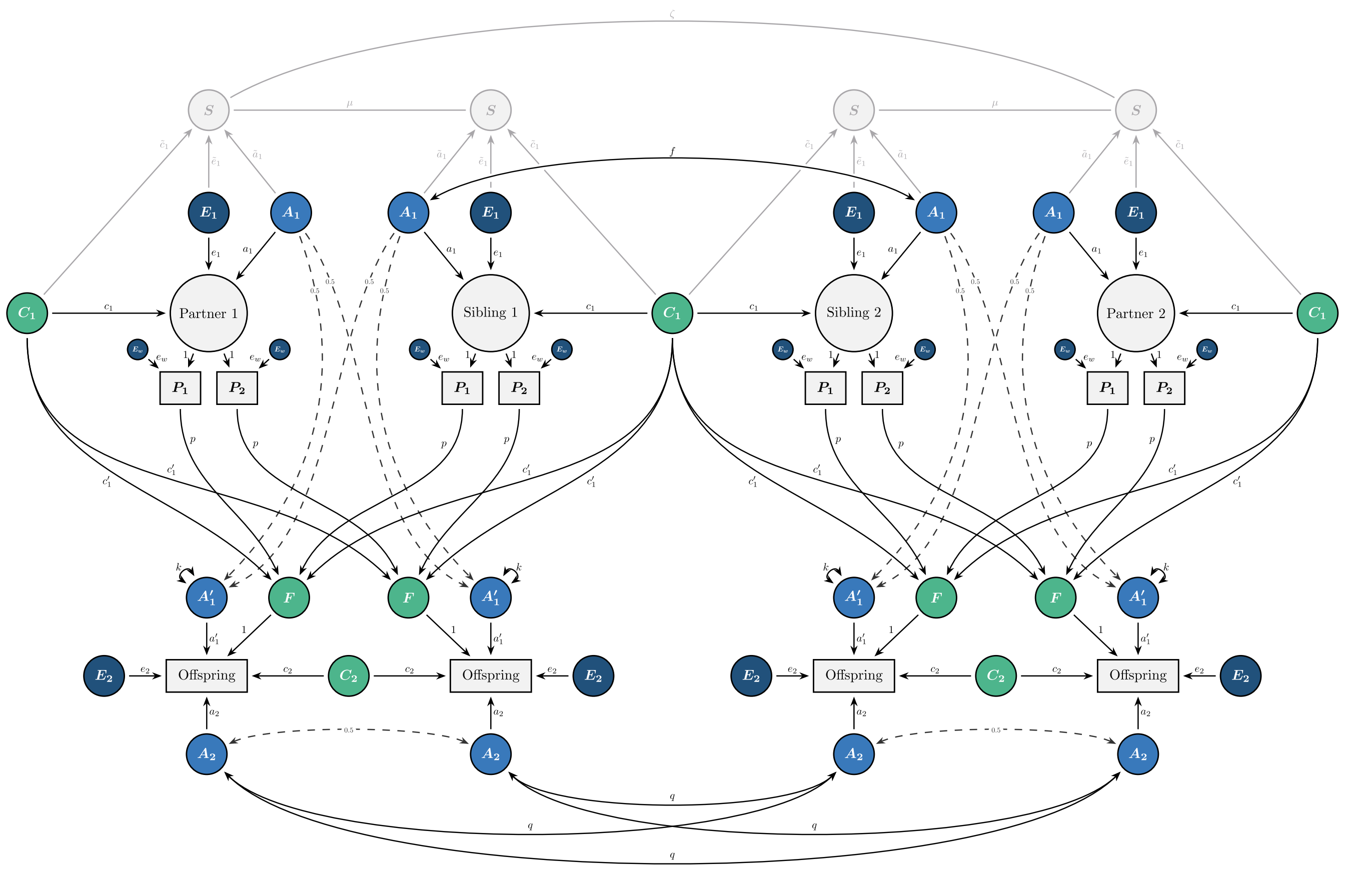


*Figure S1*: The full kinship-based model: An iAM-COTS model with variable parental phenotypes and correlated mate-preferences. For a simplified version, see Figure 1. Rectangles represent observed variables and circles represent unobserved, latent variables. All latent variables have unit variances except *F* and *Partner/Sibling*, whose variance are determined by the incoming paths. *A* = Additive Genetic Effects, *C/F* = Shared Environmental Effects, *E* = Non-Shared Environmental Effects. Because the parental phenotypes are observed multiple times, the parental E-component is split into between-person variance ($E_{1b}$) and within-person variance ($E_{1w}$). The parameters *f*, *k*, and *q* are either known or derived from other parameters. The parameter $e_2$ is constrained to 1, while the parameter $\tilde{e}_1$ is constrained to equal $e_1$. See Supplementary Table S5 for a complete overview of the parameters in the model and Supplementary Table S6 for the derived equations for the different covariances. The model is written in *OpenMx* in R and the code is available at osf.io/wq8fu/.

### Figure S2: Prevalence of any psychiatric disorder by sex and income quartile

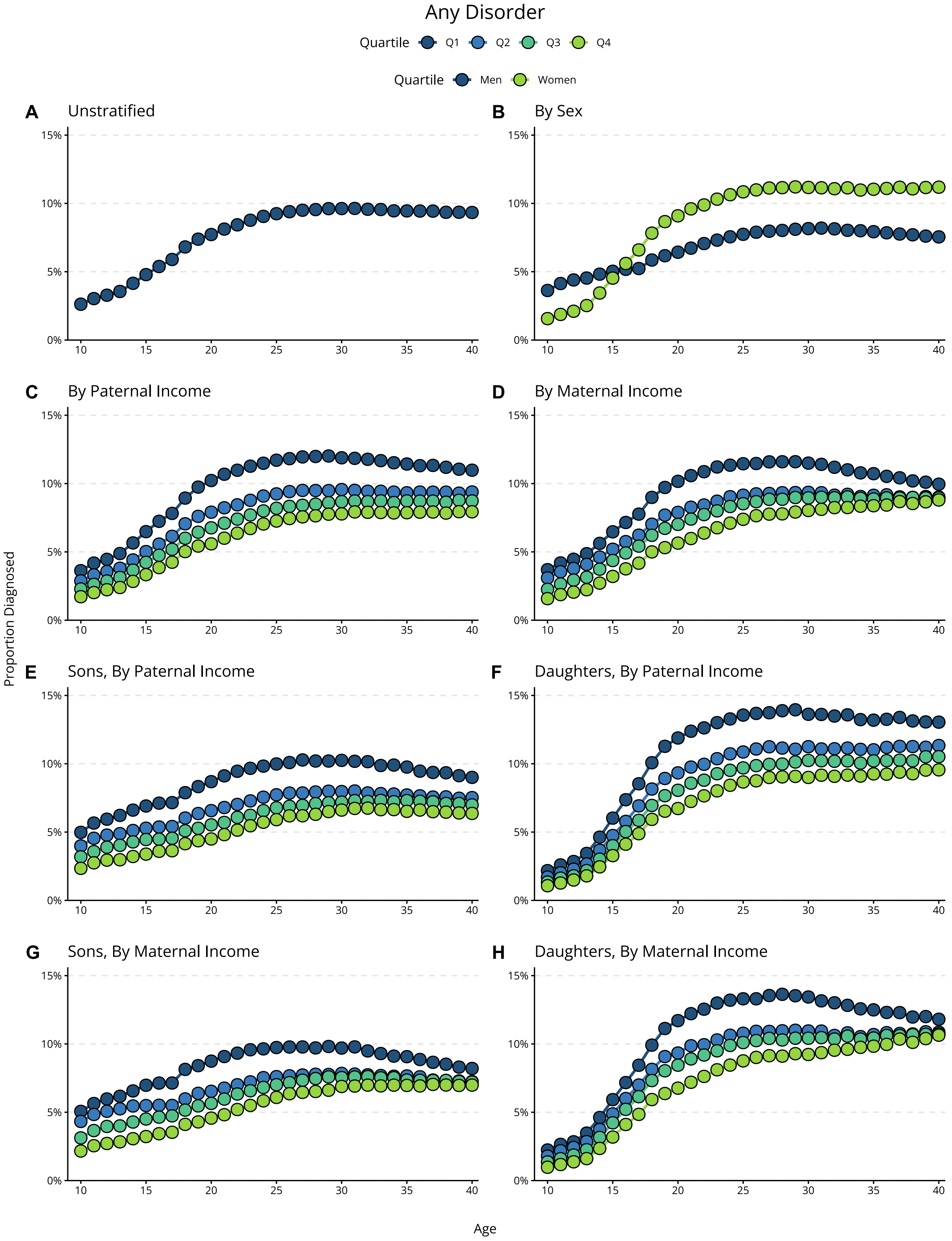


*Figure S2*: Proportion diagnosed (incl. 95% CIs) with any psychiatric disorder the year they turn a given age. Income quartiles are based on median income rank between offspring age 10 to 18. Prevalences for every diagnostic code across age, sex, and paternal/maternal income quartiles are available in an interactive figure at https://hfsu.shinyapps.io/prevalence_by_age/, or as data files at osf.io/wq8fu/.

### Figure S3: Prevalence ratios for any psychiatric disorder by sex and parental income

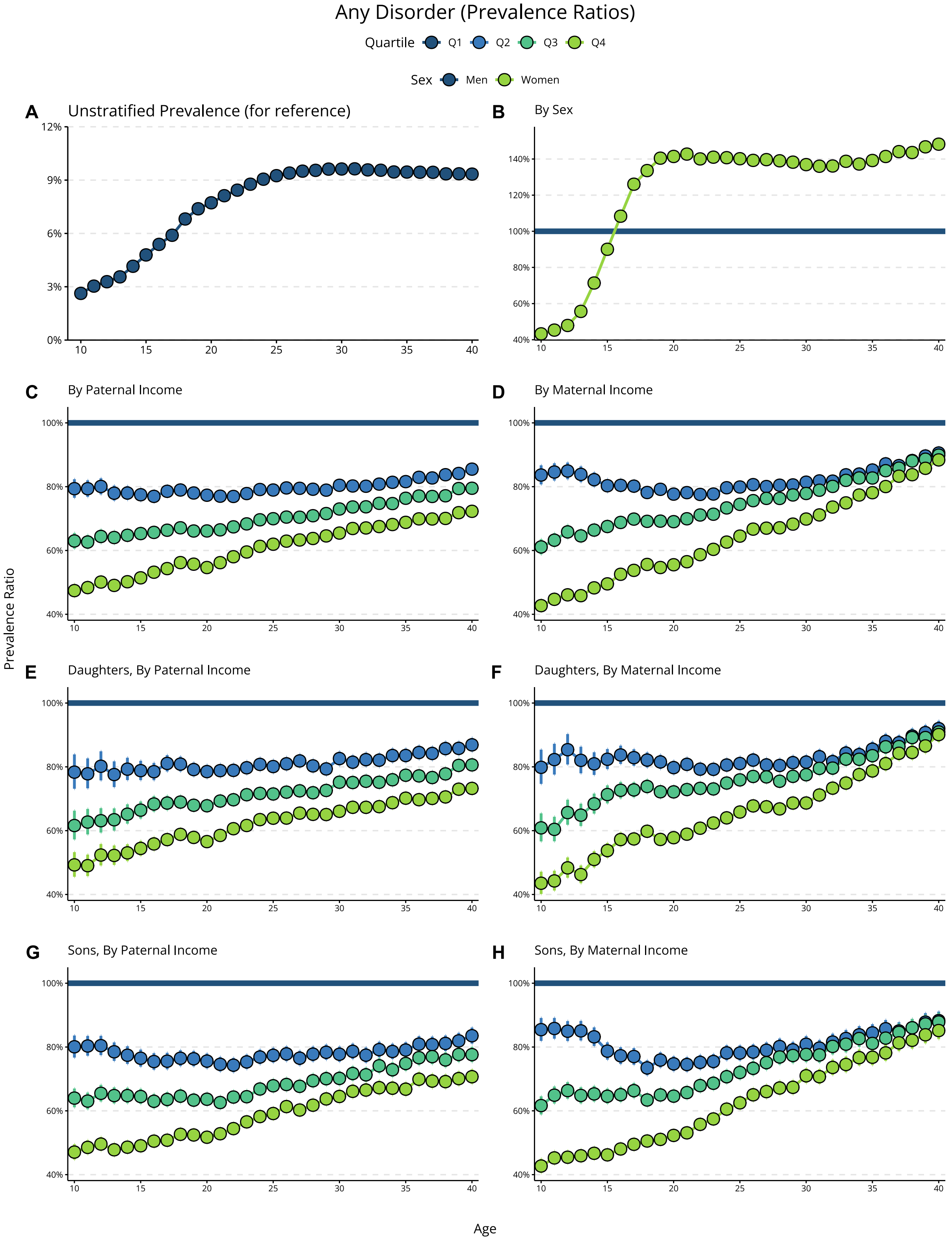


*Figure S3*: Prevalence ratios (incl. 95% CIs) for being diagnosed with any psychiatric disorder the year they turn a given age by sex and parental income. Income quartiles are based on mean income percentile between offspring age 10 to 18.

#### Figure S4: Decomposition of parent-offspring correlation when assuming direct assortment

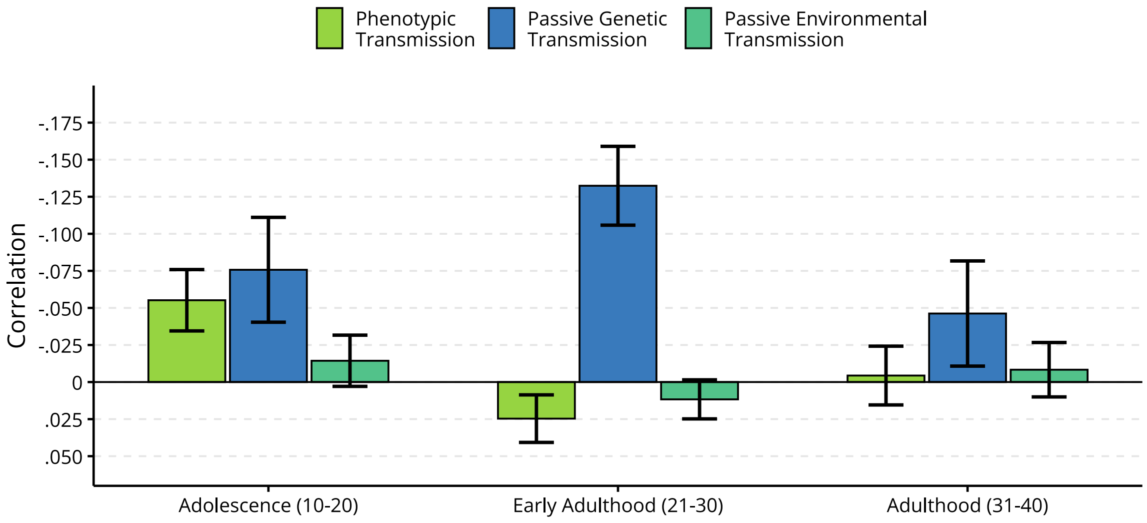


*Figure S4*: The decomposition (incl. 95% CIs) of the correlation between parental income rank and any offspring psychiatric disorders across offspring age groups ***when assuming direct assortative mating***. The correlations are assumed to reflect phenotypic transmission (i.e., direct effects of parental income rank, green), passive genetic transmission (blue), and passive environmental transmission (aquamarine). The total correlation is the sum of each component. The y-axis is reversed to make interpretation easier, so that a taller column means a larger negative correlation (lower income rank → higher risk of psychiatric disorders).

**Figure S5: Fixed-effects coefficients (any psychiatric disorder)**

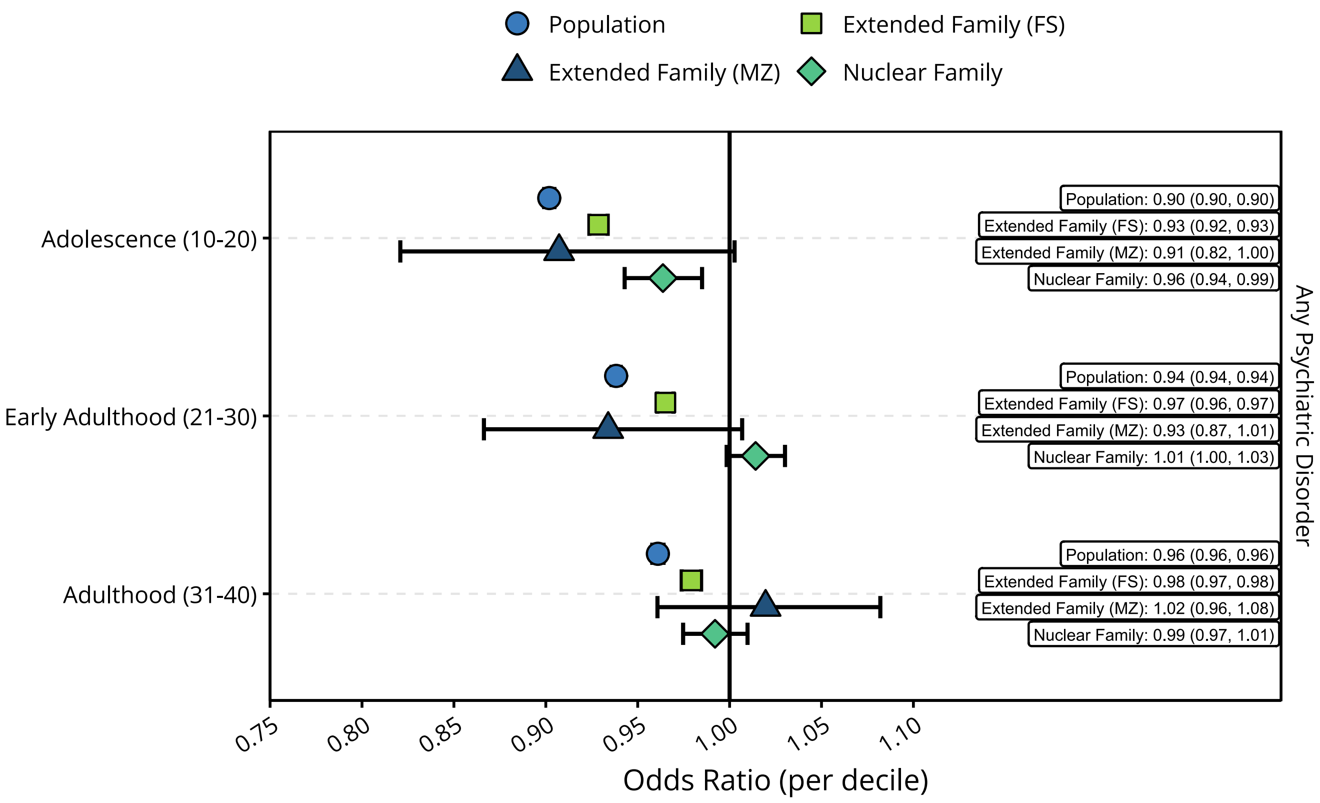


*Figure S5*: Coefficients from fixed-effects models for parental income rank on any psychiatric disorders. Coefficients are also listed in Supplementary Table S14, and in data files at osf.io/wq8fu/.

#### Figure S6: Fixed-effects coefficients (Internalizing and ADHD)

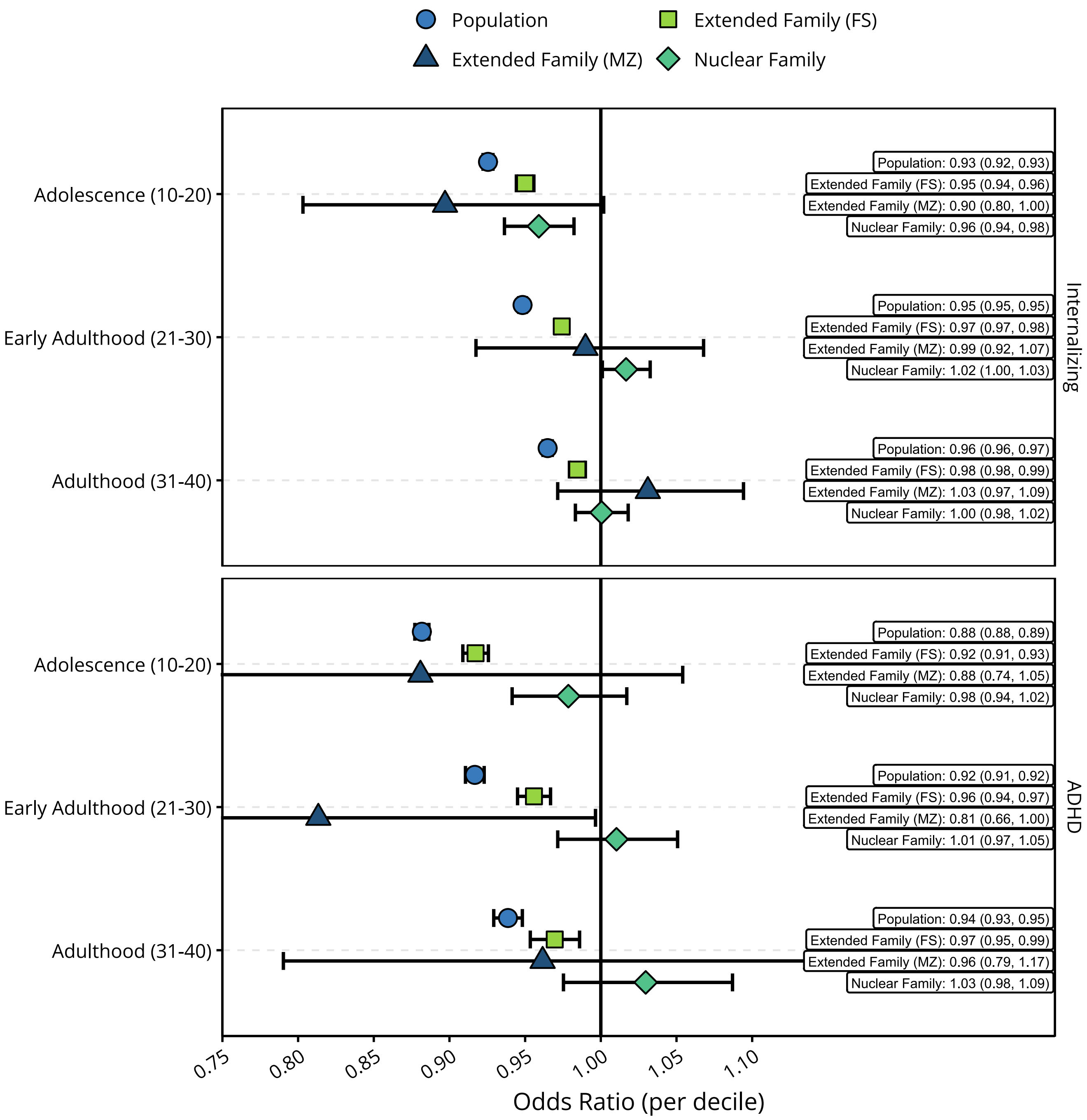


*Figure S6*: Coefficients from fixed-effects models for parental income rank on internalizing conditions (top) and ADHD (bottom). Coefficients are also listed in Supplementary Tables S15 and S16, and in data files at osf.io/wq8fu/.

**Table S1: Number of observations per age**

| Age | Number of Observations |
|---|---|
| 10 | 719,593 |
| 11 | 783,221 |
| 12 | 785,673 |
| 13 | 786,370 |
| 14 | 789,379 |
| 15 | 792,607 |
| 16 | 796,229 |
| 17 | 799,054 |
| 18 | 798,517 |
| 19 | 791,893 |
| 20 | 782,038 |
| 21 | 770,330 |
| 22 | 756,936 |
| 23 | 742,381 |
| 24 | 729,797 |
| 25 | 717,833 |
| 26 | 707,358 |
| 27 | 696,697 |
| 28 | 685,498 |
| 29 | 673,860 |
| 30 | 666,281 |
| 31 | 663,264 |
| 32 | 666,863 |
| 33 | 673,203 |
| 34 | 683,768 |
| 35 | 696,225 |
| 36 | 708,125 |
| 37 | 721,667 |
| 38 | 735,215 |
| 39 | 747,404 |
| 40 | 759,526 |

**Table S2: Number of observations per age group (kinship models)**

| | Total | | Extended Family Units | | | | | |
|---|---|---|---|---|---|---|---|---|
| **Age Group** | **Observations** | **Extended Family Units** | **Monozygotic Twins *(Male)*** | **Monozygotic Twins *(Female)*** | **Dizygotic Twins *(Male)*** | **Dizygotic Twins *(Female)*** | **Full Siblings *(Male)*** | **Full Siblings *(Female)*** |
| Adolescence (10-20) | 1,635,877 | 174,452 | 801 | 918 | 1,011 | 983 | 85,082 | 85,657 |
| Early Adulthood (21-30) | 1,453,260 | 162,875 | 890 | 915 | 1,373 | 1,226 | 77,457 | 81,014 |
| Adulthood (31-40) | 1,029,721 | 115,029 | 1,093 | 1,135 | 1,635 | 1,518 | 55,367 | 54,281 |

**Table S3: Psychological diagnostic codes in ICPC-2**

| Code | Name |
|---|---|
| P01 | Feeling anxious/nervous/tense |
| P02 | Acute stress reaction |
| P03 | Feeling depressed |
| P04 | Feeling/behaving irritable/angry |
| P05 | Senility, feeling/behaving old |
| P06 | Sleep disturbance |
| P07 | Sexual desire reduced |
| P08 | Sexual fulfilment reduced |
| P09 | Sexual preference concern |
| P10 | Stammering/stuttering/tic |
| P11 | Eating problem in child |
| P12 | Bedwetting/enuresis |
| P13 | Encopresis/bowel training problem |
| P15 | Chronic alcohol abuse |
| P16 | Acute alcohol abuse |
| P17 | Tobacco abuse |
| P18 | Medication abuse |
| P19 | Drug abuse |
| P20 | Memory disturbance |
| P22 | Child behaviour symptom/complaint |
| P23 | Adolescent behav. Symptom/complt. |
| P24 | Specific learning problem |
| P25 | Phase of life problem adult |
| P27 | Fear of mental disorder |
| P28 | Limited function/disability (p) |
| P29 | Psychological symptom/complt other |
| P70 | Dementia |
| P71 | Organic psychosis other |
| P72 | Schizophrenia |

**Table S3: Psychological diagnostic codes in ICPC-2**

| Code | Name |
|---|---|
| P73 | Affective psychosis |
| P74 | Anxiety disorder/anxiety state |
| P75 | Somatization disorder |
| P76 | Depressive disorder |
| P77 | Suicide/suicide attempt |
| P78 | Neuraesthenia/surmenage |
| P79 | Phobia/compulsive disorder |
| P80 | Personality disorder |
| P81 | Hyperkinetic disorder |
| P82 | Post-traumatic stress disorder |
| P85 | Mental retardation |
| P86 | Anorexia nervosa/bulimia |
| P98 | Psychosis NOS/other |
| P99 | Psychological disorders, other |

Prevalences for every diagnostic code across age, sex, and paternal/maternal income quartiles are available in an interactive figure at https://hfsu.shinyapps.io/prevalence_by_age/, or as data files at osf.io/wq8fu/. For the category "any psychiatric disorders", only the diagnostic codes (P70 and above) were considered.

**Table S4: Correlations in extended family**

| | | Correlation (95% CI) | | |
|---|---|---|---|---|
| **Relation** | **Zygosity[a]** | **Adolescence (10-20)** | **Early Adulthood (21-30)** | **Adulthood (31-40)** |
| ***Parent Generation (Pearson Correlations)*** | | | | |
| Within-Person | MZ | .94 *(.94, .95)* | .90 *(.89, .90)* | .91 *(.91, .92)* |
| Within-Person | DZ | .94 *(.94, .94)* | .91 *(.91, .92)* | .91 *(.90, .91)* |
| Within-Person | FS | .93 *(.93, .93)* | .91 *(.91, .91)* | .90 *(.90, .90)* |
| Partners | MZ | .18 *(.14, .22)* | .11 *(.08, .15)* | .08 *(.05, .12)* |
| Partners | DZ | .18 *(.14, .22)* | .15 *(.11, .18)* | .09 *(.06, .12)* |
| Partners | FS | .17 *(.17, .17)* | .12 *(.12, .12)* | .09 *(.09, .09)* |
| Siblings/Twins | MZ | .47 *(.43, .50)* | .45 *(.41, .49)* | .42 *(.38, .45)* |
| Siblings/Twins | DZ | .28 *(.24, .32)* | .27 *(.23, .31)* | .25 *(.21, .28)* |
| Siblings/Twins | FS | .27 *(.27, .27)* | .25 *(.24, .25)* | .24 *(.24, .25)* |
| Siblings-in-law | MZ | .18 *(.15, .22)* | .14 *(.11, .18)* | .09 *(.06, .13)* |
| Siblings-in-law | DZ | .11 *(.07, .14)* | .11 *(.08, .14)* | .09 *(.06, .12)* |
| Siblings-in-law | FS | .12 *(.11, .12)* | .09 *(.09, .10)* | .09 *(.08, .09)* |
| Co-Siblings-in-law | MZ | .18 *(.12, .25)* | .21 *(.14, .27)* | .27 *(.21, .32)* |
| Co-Siblings-in-law | DZ | .18 *(.11, .25)* | .17 *(.11, .23)* | .18 *(.13, .23)* |
| Co-Siblings-in-law | FS | .14 *(.14, .15)* | .14 *(.13, .14)* | .15 *(.14, .15)* |
| ***Intergenerational (Polyserial Correlations)*** | | | | |
| Parent-Offspring (Primary) | MZ | -.14 *(-.18, -.09)* | -.09 *(-.13, -.06)* | -.07 *(-.10, -.04)* |
| Parent-Offspring (Primary) | DZ | -.18 *(-.22, -.14)* | -.09 *(-.12, -.06)* | -.03 *(-.06, -.01)* |
| Parent-Offspring (Primary) | FS | -.15 *(-.15, -.14)* | -.10 *(-.10, -.09)* | -.06 *(-.06, -.06)* |
| Parent-Offspring (Secondary) | MZ | -.13 *(-.17, -.08)* | -.11 *(-.14, -.07)* | -.06 *(-.09, -.03)* |
| Parent-Offspring (Secondary) | DZ | -.18 *(-.22, -.13)* | -.09 *(-.12, -.05)* | -.04 *(-.06, -.01)* |
| Parent-Offspring (Secondary) | FS | -.14 *(-.15, -.14)* | -.10 *(-.10, -.10)* | -.06 *(-.06, -.06)* |
| Avuncular | MZ | -.07 *(-.13, -.02)* | -.06 *(-.10, -.01)* | -.07 *(-.11, -.03)* |
| Avuncular | DZ | -.11 *(-.17, -.06)* | -.07 *(-.11, -.03)* | -.01 *(-.05, .02)* |
| Avuncular | FS | -.07 *(-.08, -.07)* | -.05 *(-.06, -.05)* | -.04 *(-.04, -.03)* |
| Avuncular-in-law | MZ | .00 *(-.07, .07)* | -.04 *(-.10, .02)* | .00 *(-.05, .04)* |

**Table S4: Correlations in extended family**

| | | Correlation (95% CI) | | |
|---|---|---|---|---|
| **Relation** | **Zygosity**[a] | **Adolescence (10-20)** | **Early Adulthood (21-30)** | **Adulthood (31-40)** |
| Avuncular-in-law | DZ | -.05 *(-.12, .02)* | -.03 *(-.08, .02)* | -.03 *(-.07, .02)* |
| Avuncular-in-law | FS | -.06 *(-.06, -.05)* | -.03 *(-.04, -.03)* | -.02 *(-.02, -.01)* |
| ***Offspring Generation (Polychoric Correlations)*** | | | | |
| Siblings (Offspring) | MZ | .37 *(.24, .48)* | .23 *(.11, .34)* | .27 *(.18, .36)* |
| Siblings (Offspring) | DZ | .38 *(.26, .49)* | .22 *(.13, .32)* | .20 *(.11, .28)* |
| Siblings (Offspring) | FS | .32 *(.31, .33)* | .25 *(.24, .26)* | .20 *(.19, .21)* |
| Cousins (Offspring) | MZ | .02 *(-.12, .16)* | .18 *(.08, .27)* | .03 *(-.05, .12)* |
| Cousins (Offspring) | DZ | .14 *(.02, .27)* | .07 *(-.02, .16)* | .09 *(.01, .16)* |
| Cousins (Offspring) | FS | .09 *(.08, .10)* | .07 *(.06, .07)* | .06 *(.05, .07)* |

[a]MZ = Monozygotic Twins, DZ = Dizygotic Twins, FS = Full Siblings

**Table S5: Parameters in the kinship-based model**

| Description | Parameter | Free | Value |
|---|---|---|---|
| **Parent Generation** | | | |
| Additive genetic effects | $a_1$ | Free | |
| Sibling-shared Environmental effects | $c_1$ | Free | |
| Between-person unique environmental effects | $e_{1b}$ | Free | |
| Within-person effects | $e_{1w}$ | Free | |
| Assortative mating co-path | $\mu$ | Free | |
| Genetic homogamy | $\tilde{a}_1$ | Free | |
| Social homogamy | $\tilde{c}_1$ | Free | |
| Idiosyncratic homogamy | $\tilde{e}_1$ | Constrained | $\tilde{e}_1 = e_{1b}$ |
| Sibling-shared mate preferences | $\zeta$ | Free | $\zeta_{MZ} \mid \zeta_{FS}$ |
| Genetic correlation between twins/siblings | $f$ | Known/ Derived | $f_{MZ} = 1$<br>$f_{FS} = \frac{1 + \mu\tilde{a}_1^2}{2}$ |
| Parental phenotypic mean | $\bar{X}_p$ | Free | |
| **Offspring Generation** | | | |
| Additive genetic effects associated with parental phenotype | $a'_1$ | Free | |
| Direct phenotypic effects | $p$ | Free | |
| Shared environmental effects associated with parental phenotype | $c'_1$ | Free | |
| Additive genetic effects independent of parental phenotype | $a_2$ | Free | |
| Shared environmental effects independent of parental phenotype | $c_2$ | Free | |
| Unique environmental effects | $e_2$ | Constrained | $e_2 = 1$ |
| Genetic correlation between cousins | $q$ | Known | $q_{MZ} = 0.25$<br>$q_{FS} = 0.125$ |
| Recombination variance | $k$ | Derived | $k = \frac{1 - \mu\tilde{a}_1^2}{2}$ |
| Offspring phenotypic mean | $\bar{X}_o$ | Constrained | $\bar{X}_o = 0$ |
| Offspring phenotypic threshold | $\theta$ | Free | |

**Table S6: Equations implied by the iAM-COTS model in Figure S3**

| Name | Shorthand | Equation | Note |
|---|---|---|---|
| **Useful Shorthands** | | | |
| ***Variance Components in Parent generation*** | | | |
| Additive genetic variance | $V_{A1} =$ | $a_1^2$ | |
| Sibling-shared environmental variance | $V_{C1} =$ | $c_1^2$ | |
| Between-person unique environmental variance | $V_{E1b} =$ | $t_{1b}^2$ | |
| Within-person environmental variance | $V_{E1w} =$ | $e_{1w}^2$ | |
| ***Variance Components in Offspring generation*** | | | |
| Additive genetic variance associated with parental phenotype | $V_{A1P} =$ | $a'^2_1(f_{FS} + k)$ | $f_{FS} + k = 1$ |
| Family variance associated with parental phenotype | $V_F =$ | $2\sigma_p^2 p^2 + 2c'^2_1 + 4pc_1c'_1 + 2\mu(c'_1c_1 + \sigma_{PS}p)^2$ | *Both direct and passive environmental effects* |
| Additive genetic variance independent of parental phenotype | $V_{A2} =$ | $a_2^2$ | |
| Shared environmental variance independent of parental phenotype | $V_{C2} =$ | $c_2^2$ | |
| Gene-Environment covariance | $V_{rGE} =$ | $2pa_1a'_1 + 2a'_1\tilde{a}_1\mu(\tilde{c}_1c'_1 + \sigma_{PS}p)$ | |
| Unique environmental variance | $V_{E2} =$ | $e_2^2$ | |
| ***Other useful Shorthands*** | | | |
| Variance, stable parental phenotype | $\sigma_l^2 =$ | $V_{A1} + V_{C1} + V_{E1b}$ | *Variance for stable component of parental phenotype* |
| Covariance between P and S | $\sigma_{PS} =$ | $\tilde{a}_1a_1 + \tilde{c}_1c_1 + e_{1b}\tilde{e}_1$ | *Focal phenotype - Sorting factor covariance* |
| Sibling Covariance between P and S | $\delta_{PS} =$ | $f\tilde{a}_1a_1 + \tilde{c}_1c_1$ | *Covariance between focal phenotype and their siblings sorting factor* |
| Sibling Covariancen between S and S | $\delta_{SS} =$ | $f\tilde{a}_1^2 + \tilde{c}_1^2$ | *Covariance between siblings' sorting factors* |
| Genotypic correlation between full siblings | $f_{FS} =$ | $\frac{1 + \mu\tilde{a}_1^2}{2}$ | $f_{FS} = f_{DZ}$ |
| Focal Phenotype - Offspring Covariance w/o assortative mating | $\lambda_{PO} =$ | $\frac{a_1a'_1}{2} + c_1c'_1 + \sigma_p^2 p$ | *Covariance between stable parental phenotype and offspring phenotype* |

**Table S6: Equations implied by the iAM-COTS model in Figure S3**

| Name | Shorthand | Equation | Note |
|---|---|---|---|
| Sorting Factor - Offspring Covariance w/o assortative mating | $\lambda_{SO} =$ | $\frac{\tilde{a}_1 a'_1}{2} + \tilde{c}_1 c'_1 + \sigma_{PS} p$ | *Covariance between parental sorting factor and offspring phenotype* |
| **Cells in Covariance Matrix** | | | |
| ***Variances*** | | | |
| Observed Parental Phenotype | $\sigma_p^2 =$ | $\sigma_l^2 + V_{E1w}$ | |
| Offspring Phenotype | $\sigma_o^2 =$ | $V_{A1P} + V_F + V_{A2} + V_{C2} + V_{rGE} + V_{E2}$ | |
| ***Covariances in Parent Generation*** | | | |
| Within-person | | $\sigma_l^2$ | |
| Siblings/Twins | $\delta_{PP} =$ | $f V_{A1} + V_{C1}$ | |
| Partners | | $\sigma_{PS}^2 \mu$ | |
| Siblings-in-law | | $\sigma_{PS} \mu \delta_{PS}$ | |
| Co-siblings-in-law | | $\sigma_{PS}^2 (\mu^2 \delta_{SS} + \zeta)$ | |
| ***Intergenerational Covariances*** | | | |
| Parent-Offspring (Primary) | | $\lambda_{PO} + V_{E1w} p + \sigma_{PS} \mu \lambda_{SO}$ | |
| Parent-Offspring (Secondary) | | $\lambda_{PO} + \sigma_{PS} \mu \lambda_{SO}$ | *Parental phenotype for the other offspring* |
| Avuncular | $\tau =$ | $\frac{f a_1 a'_1}{2} + c_1 c'_1 + \delta_{PP} p + \delta_{PS} \mu \lambda_{SO}$ | *The genetically related uncle/aunt* |
| Avuncular-in-law | | $\sigma_{PS}(\mu((\frac{f \tilde{a}_1 a'_1}{2} + \tilde{c}_1 c'_1 + \delta_{PS} p) + \delta_{SS} \mu \lambda_{SO}) + \zeta \lambda_{SO})$ | *The non-genetically related uncle/aunt* |
| ***Covariances in Offspring Generation*** | | | |
| Siblings | | $f_{FS} a'^2_1 + \frac{V_{A2}}{2} + V_F + V_{C2} + V_{rGE}$ | |
| Cousins | | $\frac{f a'^2_1}{4} + q V_{A2} + \delta_{PP} p^2 + c'^2_1 + 2 p c_1 c'_1 + p f a_1 a'_1 + 2 \lambda_{SO} \mu (\frac{f \tilde{a}_1 a'_1}{2} + \tilde{c}_1 c'_1 + \delta_{PS} p) + \lambda_{SO}^2 (\mu^2 \delta_{SS} + \zeta)$ | |

**Table S7: Estimated Free Parameters in the Kinship-Based Model**

**Unstandardized Estimates [*Standardized Estimates*]**

| **Parameter** | **Adolescence (10-20)** | **Early Adulthood (21-30)** | **Adulthood (31-40)** | **Description** |
|---|---|---|---|---|
| $\mu$ | 0.29 [*0.87*] | 0.15 [*1.02*] | 0.00 [*1.02*] | Assortative Mating co-path |
| $\tilde{a}_1$ | 0.66 [*0.38*] | 0.64 [*0.24*] | 0.58 [*0.04*] | Genetic Homogamy *(Constrained to* $a_1$ *)* |
| $\tilde{c}_1$ | 1.60 [*0.92*] | 2.53 [*0.97*] | 14.48 [*1.00*] | Social Homogamy |
| $\tilde{e}_1$ | 0.07 [*0.04*] | 0.02 [*0.01*] | 0.00 [*0.00*] | Idiosyncratic Homogamy |
| $a_1$ | 0.66 [*0.65*] | 0.64 [*0.63*] | 0.58 [*0.58*] | Additive genetic effects |
| $c_1$ | 0.18 [*0.18*] | 0.19 [*0.19*] | 0.28 [*0.28*] | Shared environmental effects |
| $e_{1b}$ | 0.69 [*0.69*] | 0.69 [*0.68*] | 0.70 [*0.70*] | Between-person unique environmental effects |
| $e_{1w}$ | 0.26 [*0.26*] | 0.31 [*0.31*] | 0.32 [*0.32*] | Within-person effects |
| $a_1'$ | -0.27 [*-0.18*] | -0.37 [*-0.26*] | -0.21 [*-0.16*] | Additive genetic effects associated with parental phenotype |
| $p$ | -0.07 [*-0.05*] | 0.01 [*0.01*] | -0.00 [*-0.00*] | Direct phenotypic effects |
| $c_1'$ | -0.06 [*-0.04*] | -0.03 [*-0.02*] | -0.02 [*-0.02*] | Shared environmental effects associated with parental phenotype |
| $a_2$ | -1.08 [*-0.70*] | 0.91 [*0.65*] | 0.79 [*0.61*] | Additive genetic effects independent of parental phenotype |
| $c_2$ | 0.23 [*0.15*] | 0.02 [*0.02*] | 0.01 [*0.00*] | Shared environmental effects independent of parental phenotype |
| $e_2$ | 1.00 [*0.65*] | 1.00 [*0.71*] | 1.00 [*0.77*] | Unique environmental effects *(Constrained to 1)* |
| $\zeta_{FS}$ | 0.01 [-] | 0.02 [-] | 0.00 [-] | Shared mate preferences (FS) |
| $\zeta_{MZ}$ | 0.01 [-] | 0.10 [-] | 0.01 [-] | Shared mate preferences (MZ) |
| $\bar{X}_p$ | -0.02 [*-0.02*] | -0.01 [*-0.01*] | -0.00 [*-0.00*] | Parental phenotypic mean |
| $\theta$ | 1.76 [*1.15*] | 1.17 [*0.83*] | 1.09 [*0.84*] | Offspring phenotypic threshold |

**Table S8: Standardized Variance Components**

| Parameter | Adolescence (10-20) | Early Adulthood (21-30) | Adulthood (31-40) | Description |
|---|---|---|---|---|
| | **Estimates (95% CIs)** | | | |
| $V_{A1}$ | 42.6% (39.9%, 45.2%) | 40.2% (38.9%, 41.5%) | 33.3% (31.3%, 35.4%) | Additive genetic variance *(Parent Generation)* |
| $V_{C1}$ | 3.1% ( 0.2%, 6.0%) | 3.5% ( 2.0%, 5.1%) | 7.6% ( 6.6%, 8.6%) | Shared environmental variance *(Parent Generation)* |
| $V_{E1b}$ | 47.6% (45.2%, 50.1%) | 46.8% (45.7%, 48.0%) | 48.9% (47.6%, 50.2%) | Between-person unique environmental variance *(Parent Generation)* |
| $V_{E1w}$ | 6.7% ( 6.7%, 6.8%) | 9.4% ( 9.4%, 9.5%) | 10.2% (10.1%, 10.2%) | Within-person variance *(Parent Generation)* |
| $V_{A1P}$ | 3.1% ( 1.7%, 4.4%) | 6.8% ( 5.0%, 8.6%) | 2.5% ( 1.2%, 3.9%) | Additive genetic variance associated with parental phenotype *(Offspring Generation)* |
| $V_F$ | 1.4% ( 0.7%, 2.2%) | 0.1% (-0.1%, 0.3%) | 0.1% ( 0.0%, 0.3%) | Shared environmental variance associated with parental phenotype *(Offspring Generation)* |
| $V_{A2}$ | 49.3% (42.6%, 55.9%) | 42.4% (39.7%, 45.1%) | 37.5% (35.2%, 39.8%) | Additive genetic variance independent of parental phenotype *(Offspring Generation)* |
| $V_{C2}$ | 2.3% (-1.1%, 5.7%) | 0.0% (-1.0%, 1.0%) | 0.0% (-0.1%, 0.1%) | Shared environmental variance independent of parental phenotype *(Offspring Generation)* |
| $V_{rGE}$ | 1.8% ( 1.5%, 2.0%) | 0.0% (-0.4%, 0.4%) | 0.1% (-0.2%, 0.3%) | Gene-environment covariance *(Offspring Generation)* |
| $V_{E2}$ | 42.2% (38.6%, 45.8%) | 50.6% (48.6%, 52.7%) | 59.8% (57.6%, 61.9%) | Unique environmental variance *(Offspring Generation)* |

| Table S9A: Decomposition of Parent-Offspring Correlation | | | | | |
|---|---|---|---|---|---|
| | | **Estimates (95% CIs)** | | | |
| **Part[a]** | **Description** | **Adolescence (10-20)** | **Early Adulthood (21-30)** | **Adulthood (31-40)** | **Equation[b]** |
| **Total** | **Parent Offspring Correlation** | **-0.15 (-0.15, -0.14)** | **-0.10 (-0.10, -0.09)** | **-0.06 (-0.06, -0.06)** | |
| *w/o AM* | *Parent Offspring Correlation* | *-0.11 (-0.12, -0.11)* | *-0.08 (-0.08, -0.08)* | *-0.05 (-0.06, -0.05)* | |
| *AM-induced inflation* | *Parent Offspring Correlation* | *-0.03 (-0.04, -0.03)* | *-0.02 (-0.02, -0.01)* | *-0.01 (-0.01, -0.00)* | |
| **Total** | **Passive Environmental Transmission** | **-0.02 (-0.03, -0.01)** | **-0.01 (-0.02, -0.00)** | **-0.01 (-0.01, -0.01)** | |
| *w/o AM* | *Passive Environmental Transmission* | *-0.01 (-0.01, -0.00)* | *-0.00 (-0.01, -0.00)* | *-0.01 (-0.01, -0.00)* | $c_1 c'_1$ |
| *AM-induced inflation* | *Passive Environmental Transmission* | *-0.01 (-0.02, -0.01)* | *-0.01 (-0.01, -0.00)* | *-0.01 (-0.01, -0.00)* | $\sigma_{PS}\mu\tilde{c}_1 c'_1$ |
| **Total** | **Passive Genetic Transmission** | **-0.07 (-0.09, -0.05)** | **-0.09 (-0.11, -0.08)** | **-0.05 (-0.06, -0.03)** | |
| *w/o AM* | *Passive Genetic Transmission* | *-0.06 (-0.07, -0.04)* | *-0.08 (-0.09, -0.07)* | *-0.05 (-0.06, -0.03)* | $\frac{a_1 a'_1}{2}$ |
| *AM-induced inflation* | *Passive Genetic Transmission* | *-0.01 (-0.02, -0.01)* | *-0.01 (-0.02, -0.01)* | *-0.00 (-0.00, 0.00)* | $\frac{\sigma_{PS}\mu\tilde{a}_1 a'_1}{2}$ |
| **Total** | **Phenotypic Transmission** | **-0.06 (-0.07, -0.04)** | **0.01 (-0.00, 0.02)** | **-0.00 (-0.02, 0.01)** | |
| *w/o AM* | *Phenotypic Transmission* | *-0.05 (-0.06, -0.04)* | *0.01 (-0.00, 0.02)* | *-0.00 (-0.01, 0.01)* | $\sigma_p^2 p$ |
| *AM-induced inflation* | *Phenotypic Transmission* | *-0.01 (-0.01, -0.01)* | *0.00 (-0.00, 0.00)* | *-0.00 (-0.00, 0.00)* | $\mu\sigma_{PS}^2 p$ |

[a]AM: Assortative Mating. The total is the sum of the two other parts, whereas the parent-offspring correlation is the sum of the three components

[b]The equations are for the covariance, whereas the listed values are the correlations. To get the correlation, the components have been divided by the product of the parental and offspring standard deviations.

**Table S9B: Decomposition of Parent-Offspring Correlation (percentages)**

| | | Estimates (95% CIs) | | | |
|---|---|---|---|---|---|
| **Part[a]** | **Description** | **Adolescence (10-20)** | **Early Adulthood (21-30)** | **Adulthood (31-40)** | **Equation[b]** |
| **Total** | **Parent Offspring Correlation** | **100%** | **100%** | **100%** | |
| *w/o AM* | *Parent Offspring Correlation* | *76.6% ( 74.9%, 78.2%)* | *82.4% ( 80.2%, 84.6%)* | *88.8% ( 85.7%, 92.0%)* | |
| *AM-induced inflation* | *Parent Offspring Correlation* | *23.4% ( 21.8%, 25.1%)* | *17.6% ( 15.4%, 19.8%)* | *11.2% ( 8.0%, 14.3%)* | |
| **Total** | **Passive Environmental Transmission** | **13.7% ( 5.9%, 21.5%)** | **10.9% ( 2.3%, 19.4%)** | **17.6% ( 10.3%, 25.0%)** | |
| *w/o AM* | *Passive Environmental Transmission* | *4.5% ( 0.5%, 8.6%)* | *3.9% ( 0.4%, 7.4%)* | *8.4% ( 4.8%, 12.0%)* | $c_1 c'_1$ |
| *AM-induced inflation* | *Passive Environmental Transmission* | *9.1% ( 5.1%, 13.2%)* | *7.0% ( 1.9%, 12.1%)* | *9.2% ( 5.5%, 13.0%)* | $\sigma_{PS}\mu\tilde{c}_1 c'_1$ |
| **Total** | **Passive Genetic Transmission** | **47.7% ( 35.7%, 59.7%)** | **96.9% ( 83.1%, 110.8%)** | **78.6% ( 56.7%, 100.5%)** | |
| *w/o AM* | *Passive Genetic Transmission* | *39.0% ( 29.9%, 48.1%)* | *85.5% ( 74.0%, 97.0%)* | *77.0% ( 55.9%, 98.1%)* | $\frac{a_1 a'_1}{2}$ |
| *AM-induced inflation* | *Passive Genetic Transmission* | *8.7% ( 4.3%, 13.1%)* | *11.4% ( 5.9%, 16.9%)* | *1.6% ( -0.7%, 3.9%)* | $\frac{\sigma_{PS}\mu\tilde{a}_1 a'_1}{2}$ |
| **Total** | **Phenotypic Transmission** | **38.6% ( 29.0%, 48.2%)** | **-7.8% (-20.4%, 4.8%)** | **3.7% (-18.7%, 26.2%)** | |
| *w/o AM* | *Phenotypic Transmission* | *33.0% ( 24.8%, 41.2%)* | *-7.0% (-18.2%, 4.3%)* | *3.4% (-17.2%, 24.0%)* | $\sigma_p^2 p$ |
| *AM-induced inflation* | *Phenotypic Transmission* | *5.6% ( 4.2%, 7.0%)* | *-0.8% ( -2.2%, 0.5%)* | *0.3% ( -1.6%, 2.2%)* | $\mu\sigma_{PS}^2 p$ |

[a]AM: Assortative Mating. The total is the sum of the two other parts, whereas the parent-offspring correlation is the sum of the three components

[b]The equations are for the covariance, whereas the listed values are the percentages. To get the percentage, the components have been divided by the parent-offspring covariance.

**Table S10: Other derived parameters**

| | Estimates (95% CIs) | | | |
|---|---|---|---|---|
| **Parameter** | **Adolescence (10-20)** | **Early Adulthood (21-30)** | **Adulthood (31-40)** | **Equation** |
| Offspring Heritability | 52.3% (45.5%, 59.2%) | 49.2% (46.5%, 51.9%) | 40.0% (37.8%, 42.2%) | $V_{A1P} + V_{A2}$ |
| Genetic Correlation *Income and Psychiatric Disorders* | -0.24 (-0.29, -0.19) | -0.37 (-0.42, -0.33) | -0.25 (-0.32, -0.19) | $\frac{a_1 a'_1}{\sqrt{V_{A1}(V_{A1P} + V_{A2})}}$ |
| Genotypic Correlation *Full Siblings* | 0.56 (0.51, 0.62) | 0.53 (0.50, 0.56) | 0.50 (0.50, 0.50) | $f_{FS}$ |

**Table S11: Model Comparisons for Adolescence (10-20)**

| Base | Comparison | Parameters | -2LL | df | AIC | *Δ*LL | *Δ*df | *p* |
|---|---|---|---|---|---|---|---|---|
| **Full iAM-COTS-var model**[a] | | **16** | **2,619,391** | **1,612,636** | **2,619,423** | | | |
| Full iAM-COTS-var model | $c_1' = 0$ | 15 | 2,619,414 | 1,612,637 | 2,619,444 | 23.287 | 1 | $1.396 \times 10^{-6}$ |
| Full iAM-COTS-var model | $p = 0$ | 15 | 2,619,451 | 1,612,637 | 2,619,481 | 60.635 | 1 | $6.869 \times 10^{-15}$ |
| Full iAM-COTS-var model | $a_1' = 0$ | 15 | 2,619,423 | 1,612,637 | 2,619,453 | 32.706 | 1 | $1.072 \times 10^{-8}$ |
| Full iAM-COTS-var model | Direct AM | 12 | 2,623,685 | 1,612,640 | 2,623,709 | 4,293.985 | 4 | 0 |
| + Twin-shared environment | Full iAM-COTS-var model | 17 | 2,619,389 | 1,612,635 | 2,619,423 | | | |
| + Twin-shared environment | Full iAM-COTS-var model | 16 | 2,619,391 | 1,612,636 | 2,619,423 | 1.290 | 1 | $2.561 \times 10^{-1}$ |

[a]Reported parameters are from this model

| Table S12: Model Comparisons for Early Adulthood (21-30) | | | | | | | | |
|---|---|---|---|---|---|---|---|---|
| **Base** | **Comparison** | **Parameters** | **-2LL** | **df** | **AIC** | ***Δ*LL** | ***Δ*df** | ***p*** |
| **Full iAM-COTS-var model**[a] | | **16** | **2,606,467** | **1,437,978** | **2,606,499** | | | |
| Full iAM-COTS-var model | $c_1' = 0$ | 15 | 2,606,485 | 1,437,979 | 2,606,515 | 17.722 | 1 | $2.556 \times 10^{-5}$ |
| Full iAM-COTS-var model | $p = 0$ | 15 | 2,606,469 | 1,437,979 | 2,606,499 | 1.787 | 1 | $1.813 \times 10^{-1}$ |
| Full iAM-COTS-var model | $a_1' = 0$ | 15 | 2,606,617 | 1,437,979 | 2,606,647 | 149.414 | 1 | $2.328 \times 10^{-34}$ |
| Full iAM-COTS-var model | Direct AM | 12 | 2,610,006 | 1,437,982 | 2,610,030 | 3,539.318 | 4 | 0 |
| + Twin-shared environment | Full iAM-COTS-var model | 17 | 2,606,466 | 1,437,977 | 2,606,500 | | | |
| + Twin-shared environment | Full iAM-COTS-var model | 16 | 2,606,467 | 1,437,978 | 2,606,499 | 1.084 | 1 | $2.978 \times 10^{-1}$ |

[a]Reported parameters are from this model

**Table S13: Model Comparisons for Adulthood (31-40)**

| Base | Comparison | Parameters | -2LL | df | AIC | *Δ*LL | *Δ*df | *p* |
|---|---|---|---|---|---|---|---|---|
| **Full iAM-COTS-var model[a]** | | **16** | **1,870,117** | **1,024,861** | **1,870,149** | | | |
| Full iAM-COTS-var model | $c_1' = 0$ | 15 | 1,870,148 | 1,024,862 | 1,870,178 | 30.296 | 1 | $3.710 \times 10^{-8}$ |
| Full iAM-COTS-var model | $p = 0$ | 15 | 1,870,118 | 1,024,862 | 1,870,148 | 0.113 | 1 | $7.366 \times 10^{-1}$ |
| Full iAM-COTS-var model | $a_1' = 0$ | 15 | 1,870,167 | 1,024,862 | 1,870,197 | 49.719 | 1 | $1.774 \times 10^{-12}$ |
| Full iAM-COTS-var model | Direct AM | 12 | 1,873,149 | 1,024,865 | 1,873,173 | 3,031.544 | 4 | 0 |
| + Twin-shared environment | Full iAM-COTS-var model | 17 | 1,870,117 | 1,024,860 | 1,870,151 | | | |
| + Twin-shared environment | Full iAM-COTS-var model | 16 | 1,870,117 | 1,024,861 | 1,870,149 | -0.020 | 1 | $1.000 \times 10^{0}$ |

[a]Reported parameters are from this model

**Table S14 Fixed-Effects Models (Any Psychiatric Disorder)**

| Age Group | Outcome | Model | OR | 95% CI | *p* | N_Observations | N_Events |
|---|---|---|---|---|---|---|---|
| Adolescence (10-20) | Any Psychiatric Disorder | Population | 0.902 | 0.899, 0.905 | 0 | 533,958 | |
| Adolescence (10-20) | Any Psychiatric Disorder | Extended Family (FS) | 0.929 | 0.924, 0.934 | $1.598 \times 10^{-160}$ | 531,488 | 65,936 |
| Adolescence (10-20) | Any Psychiatric Disorder | Extended Family (MZ) | 0.907 | 0.821, 1.003 | $5.650 \times 10^{-2}$ | 2,470 | 274 |
| Adolescence (10-20) | Any Psychiatric Disorder | Nuclear Family | 0.964 | 0.943, 0.985 | $9.215 \times 10^{-4}$ | 533,958 | 66,210 |
| Early Adulthood (21-30) | Any Psychiatric Disorder | Population | 0.938 | 0.936, 0.941 | 0 | 475,356 | |
| Early Adulthood (21-30) | Any Psychiatric Disorder | Extended Family (FS) | 0.965 | 0.961, 0.969 | $1.736 \times 10^{-53}$ | 472,741 | 95,048 |
| Early Adulthood (21-30) | Any Psychiatric Disorder | Extended Family (MZ) | 0.934 | 0.866, 1.007 | $7.467 \times 10^{-2}$ | 2,615 | 470 |
| Early Adulthood (21-30) | Any Psychiatric Disorder | Nuclear Family | 1.014 | 0.998, 1.030 | $7.853 \times 10^{-2}$ | 475,356 | 95,518 |
| Adulthood (31-40) | Any Psychiatric Disorder | Population | 0.961 | 0.958, 0.964 | $5.561 \times 10^{-132}$ | 337,355 | |
| Adulthood (31-40) | Any Psychiatric Disorder | Extended Family (FS) | 0.979 | 0.974, 0.984 | $2.353 \times 10^{-14}$ | 333,563 | 66,554 |
| Adulthood (31-40) | Any Psychiatric Disorder | Extended Family (MZ) | 1.020 | 0.961, 1.082 | $5.225 \times 10^{-1}$ | 3,792 | 727 |
| Adulthood (31-40) | Any Psychiatric Disorder | Nuclear Family | 0.992 | 0.975, 1.010 | $3.775 \times 10^{-1}$ | 337,355 | 67,281 |

**Table S15 Fixed-Effects Models (Internalizing Conditions)**

| Age Group | Outcome | Model | OR | 95% CI | *p* | N_Observations | N_Events |
|---|---|---|---|---|---|---|---|
| Adolescence (10-20) | Internalizing | Population | 0.925 | 0.922, 0.929 | 0 | 533,958 | |
| Adolescence (10-20) | Internalizing | Extended Family (FS) | 0.950 | 0.944, 0.956 | $1.979 \times 10^{-61}$ | 531,488 | 47,683 |
| Adolescence (10-20) | Internalizing | Extended Family (MZ) | 0.897 | 0.803, 1.002 | $5.406 \times 10^{-2}$ | 2,470 | 198 |
| Adolescence (10-20) | Internalizing | Nuclear Family | 0.959 | 0.936, 0.982 | $6.201 \times 10^{-4}$ | 533,958 | 47,881 |
| Early Adulthood (21-30) | Internalizing | Population | 0.948 | 0.946, 0.951 | 0 | 475,356 | |
| Early Adulthood (21-30) | Internalizing | Extended Family (FS) | 0.974 | 0.970, 0.979 | $1.818 \times 10^{-29}$ | 472,741 | 92,964 |
| Early Adulthood (21-30) | Internalizing | Extended Family (MZ) | 0.990 | 0.918, 1.068 | $7.923 \times 10^{-1}$ | 2,615 | 451 |
| Early Adulthood (21-30) | Internalizing | Nuclear Family | 1.017 | 1.001, 1.033 | $3.688 \times 10^{-2}$ | 475,356 | 93,415 |
| Adulthood (31-40) | Internalizing | Population | 0.965 | 0.962, 0.968 | $2.143 \times 10^{-109}$ | 337,355 | |
| Adulthood (31-40) | Internalizing | Extended Family (FS) | 0.985 | 0.979, 0.990 | $1.052 \times 10^{-8}$ | 333,563 | 68,337 |
| Adulthood (31-40) | Internalizing | Extended Family (MZ) | 1.031 | 0.972, 1.094 | $3.135 \times 10^{-1}$ | 3,792 | 761 |
| Adulthood (31-40) | Internalizing | Nuclear Family | 1.000 | 0.983, 1.018 | $9.561 \times 10^{-1}$ | 337,355 | 69,098 |

| **Table S16 Fixed-Effects Models (ADHD)** | | | | | | | |
|---|---|---|---|---|---|---|---|
| **Age Group** | **Outcome** | **Model** | **OR** | **95% CI** | ***p*** | **N_Observations** | **N_Events** |
| Adolescence (10-20) | ADHD | Population | 0.882 | 0.877, 0.886 | 0 | 533,958 | |
| Adolescence (10-20) | ADHD | Extended Family (FS) | 0.917 | 0.909, 0.926 | $1.734 \times 10^{-75}$ | 531,488 | 20,904 |
| Adolescence (10-20) | ADHD | Extended Family (MZ) | 0.881 | 0.736, 1.054 | $1.661 \times 10^{-1}$ | 2,470 | 85 |
| Adolescence (10-20) | ADHD | Nuclear Family | 0.979 | 0.941, 1.017 | $2.720 \times 10^{-1}$ | 533,958 | 20,989 |
| Early Adulthood (21-30) | ADHD | Population | 0.917 | 0.911, 0.923 | $3.012 \times 10^{-141}$ | 475,356 | |
| Early Adulthood (21-30) | ADHD | Extended Family (FS) | 0.956 | 0.945, 0.967 | $7.936 \times 10^{-15}$ | 472,741 | 12,309 |
| Early Adulthood (21-30) | ADHD | Extended Family (MZ) | 0.813 | 0.664, 0.996 | $4.615 \times 10^{-2}$ | 2,615 | 57 |
| Early Adulthood (21-30) | ADHD | Nuclear Family | 1.010 | 0.972, 1.051 | $6.066 \times 10^{-1}$ | 475,356 | 12,366 |
| Adulthood (31-40) | ADHD | Population | 0.939 | 0.929, 0.948 | $6.541 \times 10^{-35}$ | 337,355 | |
| Adulthood (31-40) | ADHD | Extended Family (FS) | 0.970 | 0.953, 0.986 | $3.045 \times 10^{-4}$ | 333,563 | 5,581 |
| Adulthood (31-40) | ADHD | Extended Family (MZ) | 0.962 | 0.790, 1.170 | $6.952 \times 10^{-1}$ | 3,792 | 45 |
| Adulthood (31-40) | ADHD | Nuclear Family | 1.030 | 0.975, 1.087 | $2.908 \times 10^{-1}$ | 337,355 | 5,626 |
